# Emergency-granulopoiesis trajectory, but not the CD4/NK lymphocyte trajectory, is directionally reproduced as a correlate of sepsis mortality: a cross-cohort transcriptomic study with external testing and clinical-analogue triangulation

**DOI:** 10.64898/2026.08.04.26359671

**Authors:** Lei Su, Lin Zhang, Wencai Huang, Chunmei Gui, Fang Gong

## Abstract

In early sepsis the direction in which blood immune-cell transcriptional programmes move may carry prognostic information beyond a single baseline measurement, but whether such trajectory associations survive independent testing is unknown. We scored five immune modules, frozen before analysis, in three public longitudinal whole-blood microarray sepsis cohorts and fitted a logistic model ladder fixed in advance to the change per 24 hours in the two cohorts with mortality data (82 patients, 24 deaths), pooling by inverse-variance fixed-effect meta-analysis with Benjamini-Hochberg control. No association survived correction for multiple testing. The two leading signals were a rising CD4/NK lymphocyte trajectory associated with lower mortality (pooled odds ratio 0.53, 95% confidence interval 0.31 to 0.90) and a rising emergency-granulopoiesis trajectory associated with higher mortality (1.60, 0.92 to 2.79), both per one standard deviation. We then tested both in an independent transcriptomic cohort with serial sampling (63 patients, 15 deaths), scored by the identical frozen method, and against their cell-count analogues in an intensive-care database of 12,607 adults meeting Sepsis-3 criteria, of whom 744 to 4,206 had the serial measurements each analogue required. Independent testing separated the two signals, in the order opposite to the one discovery had suggested. The emergency-granulopoiesis association was reproduced in direction and effect size without reaching conventional significance on its own (validation odds ratio 1.72, 0.92 to 3.20, p=0.088; pooled 1.65, 1.09 to 2.50), was positive in all nine sensitivity analyses, each fixed before the estimates were examined, and was supported by two of its three analogues, including the neutrophil-to-lymphocyte ratio (1.31, 1.20 to 1.42). The CD4/NK association did not reproduce (1.06, 0.59 to 1.89), was null in the window most favourable to it, and received no support from an analogue well powered to detect the discovery effect. The discovery signal that looked most consistent failed independent testing.

## 1. Introduction

Sepsis is a life-threatening organ dysfunction caused by a dysregulated host response to infection, and it remains a leading cause of in-hospital and intensive-care death worldwide. A central feature of the septic host response is a rapidly evolving mixture of pro-inflammatory activation and immunosuppression that varies markedly between patients and over time [1, 2]. Whole-blood transcriptomics has become a powerful lens on this heterogeneity, and successive studies have defined reproducible molecular subgroups—sepsis response signatures, molecular endotypes, and consensus transcriptomic subtypes—that stratify patients by immune-dysfunction severity and prognosis [3, 4, 5]. These frameworks have advanced the field from single-marker thinking toward multi-gene programs that capture coordinated immune states.

*Most transcriptomic stratification, however, has been anchored on a single time point— typically at or near intensive-care admission [3, 6]. Sepsis is intrinsically dynamic: the same admission profile may resolve or deteriorate over subsequent days, and it is increasingly recognized that how the immune state changes may be as informative as where it starts. Serial cellular studies support this view. Monocyte HLA-DR expression, a canonical marker of sepsis-induced immunoparalysis, discriminates outcome more clearly by its trajectory—the decline from early to later time points is steeper in non-survivors—than by any single reading [7, 8]. Likewise, sepsis induces an early, profound loss of CD4 T-cell and natural-killer (NK)-cell numbers and function, and recovery of these compartments over time is associated with improved survival [1, 9, 10], including primary longitudinal evidence linking lymphocyte recovery to lower mortality [11]. Longitudinal multi-marker studies have begun to formalize this by clustering immune-marker trajectories into distinct immunotypes with divergent outcomes [12]; complementary large cross-sectional transcriptomic analyses corroborate immunosuppression-associated subgroups early in sepsis [13]. Two recent studies come closer still: serial whole-blood transcriptomes have been profiled across the first week of sepsis and the resulting signature tested against several external mortality cohorts by single-sample enrichment scoring [1], and latent-class modelling of serial lymphocyte counts in more than two thousand patients has identified trajectory phenotypes that separate 28-day mortality, with external confirmation in MIMIC-IV [33]. Neither asks whether change adds to baseline for a module fixed in advance. Yet direct, prespecified tests of whether the slope or direction of change of defined immune programs improves outcome prediction beyond baseline remain uncommon, and are frequently under- powered and under-reported*.

Three cell-type programs recur across this literature as candidates whose dynamics might matter. First, emergency granulopoiesis—the stress-driven expansion of immature, often immunosuppressive neutrophils—characterizes the most severe transcriptomic endotype and associates with worse outcomes [14, 15], although circulating immature-granulocyte fractions track severity more reliably than mortality [16]. Second, monocyte MHC-II (HLA-DR) suppression marks immunoparalysis and predicts death and secondary infection [8, 17]. Third, lymphocyte and NK-cell dysfunction, and the capacity to recover it, shape immune-recovery dynamics and later clinical outcomes in sepsis [9, 18]; failure of lymphocyte recovery is associated with higher longer-term mortality [11, 19]. The interferon response adds a further, more variable axis [20].

*Here we address a focused question: during early sepsis, does the within-patient rate and direction of change in defined immune-module activity relate to mortality beyond baseline immune state alone, and does any such relationship survive independent testing? We first conducted an exploratory cross-cohort analysis of three longitudinal whole-blood sepsis datasets under a written preregistered plan using five a-priori immune modules and a logistic model ladder fixed before the analysis was run. Because the discovery cohorts are small and no association passed multiplicity correction, we then tested the two leading signals twice: in an independent longitudinal transcriptomic cohort using the identical frozen modules and analysis, and against their routine cell-count analogues in a large intensive-care database. The entire analysis is released as a frozen, checksum-verified, end-to-end reproducible package*.

## 2. Methods

### 2.1 Study design and cohorts

This was a retrospective, exploratory, cross-cohort reanalysis of publicly available longitudinal whole-blood transcriptomic sepsis datasets, conducted under a written analysis plan finalised before any outcome model was run. The plan fixed the modules, exposures, outcomes, model ladder, covariate budget, multiplicity family and sensitivity analyses in advance. It was an internal written preregistration and was not deposited in a public registry, so it carries no independent timestamp; the document is provided in full as supporting information, together with an itemised log of every difference between it and the analysis reported here. Because those differences are numerous and some are material — among them the primary readout, the omission of the planned demographic covariates, the use of a fixed-effect rather than a random-effects primary pooling estimator, the exclusion of M5 from the confirmatory family, the change in the M5 construct, and the non-execution of the planned landmark and early-death sensitivity arms — the study is best described as a retrospective exploratory analysis guided by a written pre-analysis plan, with all material deviations documented, rather than as a preregistered confirmatory analysis. Two dated resolution notes concerning the GSE54514 outcome horizon were added to the plan before manuscript preparation and are marked as such within it. Three Gene Expression Omnibus (GEO) series were included (Table 1; Supplementary Methods):

- **GSE95233** — 51 patients with serial sampling; day-28 mortality confirmed from the GEO series record and its linked BioProject (PRJNA376553); 17 non-survivors and 34 survivors. Sampling was two time points per patient by design, permitting a change-per-24 h exposure but not a multi-time-point slope.
- **GSE54514 — 36 patients; outcome recorded as survivor vs non-survivor (“reported mortality”; exact follow-up horizon not stated in the accessible source). After applying the eligibility and fitting rules fixed before the analysis was run (a day-1 baseline and at least two serial samples for trajectory eligibility; at least eight observations and both outcome classes present for a model to be fitted), 31 patients (7 events) entered change models; 29 patients with ≥3 serial samples (7 events) supported a secondary ordinary-least-squares (OLS) slope exposure. Illness severity was available as APACHE II (range 9–35).**
- **GSE57065** — 28 patients, of whom 26 had complete H00/H24/H48 sampling triplets; **no mortality outcome was collected**. This cohort was used only for early immune-state dynamics and associations with binary SAPS-II severity (12 high, 14 low), and contributed no mortality models.

**Table 1.** Cohort characteristics and descriptive module dynamics by outcome. Source: ‘table1_descriptive_by_outcome.csv’ (per-cohort baseline and raw-change module means ± SD by survivor/non-survivor, with Mann–Whitney p-values) and cohort metadata (Section 2.1). Rendered in full in the typeset manuscript.

| cohort | module | module_label | n_surv | n_nonsurv | baseline_surv_mean | baseline_surv_sd | baseline_nonsurv_mean | baseline_nonsurv_sd | baseline_MW_p | rawchange_surv_mean | rawchange_surv_sd | rawchange_nonsurv_mean | rawchange_nonsurv_sd | rawchange_MW_p |
| --- | --- | --- | --- | --- | --- | --- | --- | --- | --- | --- | --- | --- | --- | --- |
| GSE 95233 | M1_EG_immature_neutrophil | M1 Emergency granulopoiesis | 34 | 17 | 0.425 | 0.356 | 0.629 | 0.342 | 0.0617 | -0.265 | 0.466 | -0.089 | 0.319 | 0.0564 |
| GSE 95233 | M2_monocyte_MHCII_suppression | M2 Monocyte MHC-II suppression | 34 | 17 | -0.229 | 0.789 | -0.713 | 0.839 | 0.0564 | 0.372 | 0.714 | -0.008 | 0.85 | 0.1122 |
| GSE 95233 | M3_CD4_NK_dysfunction | M3 CD4/NK lymphocyte | 34 | 17 | -0.322 | 0.717 | -0.407 | 0.806 | 0.7568 | 0.3 | 0.704 | -0.284 | 0.938 | 0.0287 |
| GSE 95233 | M4_interferon | M4 Interferon | 34 | 17 | -0.074 | 1.11 | -0.289 | 1.085 | 0.5033 | -0.008 | 0.664 | 0.064 | 0.519 | 0.8338 |
| GSE 54514 | M1_EG_immature_neutrophil | M1 Emergency granulopoiesis | 24 | 7 | 0.016 | 1.101 | -0.262 | 0.794 | 0.4443 | 0.082 | 1.356 | 1.444 | 0.786 | 0.0124 |
| GSE 54514 | M2_monocyte_MHCII_suppression | M2 Monocyte MHC-II suppression | 24 | 7 | -0.194 | 0.968 | -1.165 | 0.875 | 0.0256 | 0.297 | 1.441 | 1.688 | 1.646 | 0.0333 |
| GSE 54514 | M3_CD4_NK_dysfunction | M3 CD4/NK lymphocyte | 24 | 7 | -0.554 | 1.218 | -0.089 | 1.06 | 0.417 | 0.646 | 1.64 | -0.361 | 1.484 | 0.2342 |
| GSE 54514 | M4_interferon | M4 Interferon | 24 | 7 | -0.378 | 1.188 | 0.545 | 0.937 | 0.0847 | 0.114 | 1.299 | 0.357 | 0.852 | 0.6945 |

Raw data, cohort definitions, and outcome definitions were fixed at the frozen stage and are not modified in this report.

### 2.2 Immune modules and single-sample scoring

Five a-priori immune modules were defined from the literature and frozen before scoring (gene membership in ‘modules_frozen.json’; Supplementary Methods): M1 emergency granulopoiesis / immature neutrophils (21 genes; higher = worse; the score is a whole-blood transcriptional correlate of emergency granulopoiesis and is not a direct measurement of the marrow process, so it is more precisely read as emergency-granulopoiesis-associated transcriptional activity, and like every bulk-blood module score it cannot be separated into a change in cell composition and a change in per-cell transcription), M2 monocyte MHC-II suppression (10 genes; the score is oriented to the antigen-presentation programme itself, so higher values indicate greater MHC-II/antigen-presentation activity and lower values indicate greater suppression), M3 CD4/NK lymphocyte-associated transcriptional activity (15 genes: CD3D, CD3E, IL7R, CD247, KLRB1, GZMB, GZMK, PRF1, NKG7, KLRD1, GNLY, CCL5, IL2RB and the inhibitory-receptor genes PDCD1 and LAG3; higher values indicate higher within-sample relative expression of the module genes, and the functional direction is not uniquely determined), M4 interferon response (17 genes), and M5 innate-inflammatory transcriptional activity (12 genes; the written plan had defined this module as a contrast between persistent innate inflammation and a recovery/adaptive axis, but the implemented score contains only same-direction innate-inflammatory genes and no recovery or adaptive genes, so it measures innate-inflammatory transcriptional activity and is named accordingly here rather than as inflammation-versus-recovery; it was designated exploratory before the analysis was run and excluded from the confirmatory family and from pooling, although twelve of its thirteen reference genes map to both platforms and it is scored, fitted and reported for every cohort. Both the construct change and the exclusion are itemised in the deviation log). Per-sample module scores were computed as within-sample rank-percentile means: within each sample, genes were ranked by expression, converted to percentiles, and averaged over the module’s usable genes. Scores were frozen at this point; these frozen module scores are inputs and are not recomputed or altered here. Single-sample gene-set enrichment (ssGSEA) scores were computed on the identical frozen gene lists and are reported as the alternative-scoring sensitivity analysis. Single-sample module scoring is an established approach for immune-signature quantification [21, 22, 23, 24] but has recognized sensitivity to platform, normalization, and sparsity, as characterized in method evaluations including two preprints [25, 26] (preprints; not peer-reviewed), which we address in the Limitations.

Direction and interpretation of M3. The preregistered plan labelled M3 as ‘CD4 T-cell and NK-cell dysfunction’ and hypothesised that sustained dysfunction would be associated with worse outcome, but it did not specify the numerical orientation of the score. In the implemented score, all 15 genes contributed in the same direction to an unweighted within-sample rank-percentile mean. Because most constituents are lymphocyte/NK identity or cytotoxic-effector transcripts, higher values operationally indicate greater CD4/NK-associated transcriptional activity. However, the module also contains the inhibitory-receptor genes PDCD1 and LAG3 and was measured in bulk whole blood; it therefore cannot distinguish lymphocyte abundance, effector activity and exhaustion, and should not be equated with preserved lymphocyte function. We consequently use the direction-neutral label ‘M3 CD4/NK lymphocyte-associated transcriptional activity’. This interpretive reannotation was not prespecified and is declared in

S8 Appendix. The stored variable name remains M3_CD4_NK_dysfunction in the archived files, as a legacy identifier that does not carry the final biological characterisation. Associations of increasing M3 with mortality are reported as observed exploratory directions rather than as confirmation of an a-priori protective or recovery hypothesis; the corresponding change association did not survive Benjamini– Hochberg FDR correction (meta q=0.075; Section 3.3).

### 2.3 Exposures and outcome model ladder

The binary outcome was in-hospital/short-term mortality (survivor vs non-survivor), coded identically across mortality cohorts; because coding is binary, the analysis is horizon-independent and no model was rerun for the unresolved GSE54514 horizon.

For each cohort × module, three within-cohort standardized exposures were defined once, over the appropriate modeled subset: baseline (first available sample, z-scored within cohort × module over the trajectory-primary set); change-per-24 h (first-to-last change divided by elapsed time, z-scored); and, where ≥3 serial samples existed (GSE54514 only), an OLS slope (per-patient linear slope over time, z-scored over the slope-eligible set). All standardization was performed once in the analysis driver so that every downstream model consumed identical, precomputed z-scores.

A logistic model ladder, fixed before the analysis was run, was fit one module at a time (no clinical covariate in the primary models, given event constraints; see 2.4). The rungs are numbered Model 0 to Model 3 throughout, to keep them distinct from the immune modules M1–M5; the frozen result tables retain their original identifiers (‘M0_intercept’, ‘M1_baselinè, ‘M2_changè, ‘M3_both’):

- Model 0 (null / intercept-only): reference model; reported for every module to anchor the ladder (AIC = −2·log-likelihood + 2; apparent AUC 0.5; no effect estimate).
- Model 1 (baseline): mortality ∼ baseline module z-score.
- Model 2 (change): mortality ∼ change-per-24 h (or OLS slope) z-score.
- Model 3 (baseline + change): mortality ∼ baseline + change.

Odds ratios (OR) are reported per +1 standard deviation of the exposure, with Wald 95% confidence intervals, log-odds, standard errors, and p-values taken directly from the maximum-likelihood fit. The incremental value of trajectory over baseline—the study’s primary question—was assessed by likelihood-ratio tests (LRT) comparing nested models (Model 3 vs Model 1, Model 3 vs Model 2) and by Akaike information criterion (AIC).

### 2.4 Feasibility, separation, and covariate policy

*Event-per-variable (EPV) constraints were explicit: GSE95233 supported EPV about 17 (17 events, one covariate), whereas GSE54514 supported EPV about 3.5, permitting only a single exposure per model. APACHE II adjustment in GSE54514 was therefore prespecified as an over-parameterized sensitivity analysis, not a primary model. Complete or quasi-complete separation was monitored for every fit and was measured directly during bootstrap resampling. Maximum-likelihood estimates converged for all reported point estimates, and a Firth penalized-likelihood fallback was available. Under separation, ordinary maximum-likelihood odds ratios and their intervals can be unstable, and penalized-likelihood approaches are the recommended remedy [27, 28, 29, 30]; the observed incidence of separation and its consequences are reported in Section 3.8*.

### 2.5 Cross-cohort meta-analysis and multiple-testing control

Per-cohort log-odds and standard errors for the change-per-24 h (Model 2) exposure were pooled across the two mortality cohorts by inverse-variance fixed-effect meta-analysis (primary), with a DerSimonian–Laird random-effects estimate reported as a secondary/exploratory sensitivity; between-cohort heterogeneity was summarized by Cochran’s Q, I², and τ². Meta-analysis was restricted to the four primary modules M1–M4; M5 was excluded from pooling and from the confirmatory family as an exploratory module, a decision taken by the analysts at the frozen stage rather than one specified in the plan. Its estimates are reported in full in the supporting information.

Severity-only cohort analyses (GSE57065). This cohort has no mortality outcome and was analysed only for early dynamics. Analyses were restricted to the 26 patients with complete H00/H24/H48 sampling. Module scores were modelled by linear mixed-effects regression with a random intercept per patient and maximum-likelihood estimation, with time measured in days from the first sample. Two models were fitted for each module and are reported separately: a pooled time-trend model containing time alone, from which the overall early trajectory is read; and a model containing time, binary SAPS-II stratum and their interaction, from which the severity-modification of that trajectory is read. Because the time coefficient in the interaction model is the slope within the low-severity stratum rather than the pooled slope, the two must not be read from the same fit. Early-change (H00 to H24) shifts were compared between severity strata by Mann-Whitney test with Benjamini-Hochberg correction.

Multiplicity was controlled with the Benjamini–Hochberg (BH) FDR procedure. The confirmatory family, fixed before the analysis was run, was M1–M4 × {baseline (Model 1), change (Model 2)} within each mortality cohort. The preregistered plan had placed all five modules in this family; restricting it to M1–M4 was an analyst decision taken at the frozen stage and is itemised in the deviation log, and it makes the correction less stringent than planned; the slope analysis and the meta-analysis were handled as separate families. An association was considered confirmatory only if it achieved q<0.05 within its family; the q<0.05 threshold itself is preregistered; all other signals are reported as suggestive or null. The meta-analysis method, the BH procedure, and the family definitions were fixed at the frozen stage and are unchanged here. One further sensitivity analysis was added at pre-submission review and is not part of the frozen specification: a landmark analysis at 48 h, restricted to patients whose last frozen sample falls at 48 h, so that every included patient was demonstrably alive at the landmark and no exposure information post-dates it. Exposures were re-standardised within the landmark set, and the change-exposure model was refitted per cohort and pooled by the same inverse-variance fixed-effect method (S2d Table).

### 2.6 Discrimination, calibration, and bootstrap diagnostics

Apparent (in-sample) discrimination was summarized by the area under the receiver-operating-characteristic curve (AUC). Optimism-corrected discrimination used Harrell’s bootstrap optimism correction (B=1000 resamples, fixed seed), with a stability rule, fixed before the analysis was run, requiring at least 50% valid resamples; calibration was summarized by an in-sample logistic recalibration (calibration slope and intercept). For every bootstrap fit we recorded whether the software raised a separation warning, whether the fit converged, and whether the linear predictor still yielded a finite AUC, so that the occurrence of separation could be distinguished from its consequences.

An earlier version of this pipeline reported the optimism-corrected AUC as not estimable for any model, attributing the absence of valid bootstrap replicates to complete or quasi-complete separation. On audit that attribution was incorrect: the missing values arose from a programming defect in the bootstrap helper, not from separation. The defect and its correction are documented in full in a companion methodological report and in the public code archive. After correction, optimism-corrected AUC was estimable for every model containing at least one exposure, and separation was measured directly rather than assumed. All discrimination values reported here come from the corrected computation.

The calibration slope and intercept, by contrast, derive from an in-sample logistic recalibration and do not use the bootstrap; they are estimable and reproducible (slope ≈ 1.0, intercept ≈ 0 by construction) and are reported, but must be interpreted as apparent / in-sample calibration only. True calibration requires an external validation cohort, which does not exist here.

### 2.7 External testing and clinical-analogue triangulation

Two independent validation analyses were added after the discovery analysis was frozen. Both used the same frozen module definitions, the same scoring method, and the same exposure and model definitions as the discovery analysis; no parameter was tuned to the validation data.

Transcriptomic validation (same modality). We used the GAinS cohort (ArrayExpress E-MTAB-5273; Burnham and colleagues), an independent prospective study of adults admitted to UK intensive care units with sepsis due to community-acquired pneumonia or faecal peritonitis, profiled on the Illumina HumanHT-12 v4 array. We used the authors’ own variance-stabilised normalised matrix (27,159 probes, 231 samples) exactly as deposited, without renormalisation. Probe identifiers were mapped to gene symbols through the GPL10558 platform annotation (100% of probes mapped; 21,544 genes after collapsing to the highest-expressing probe per gene). The five frozen modules were scored by the same within-sample rank-percentile method used in discovery; module gene recovery was 21/21 (M1), 10/10 (M2), 14/15 (M3), 16/17 (M4) and 12/12 (M5). Serial sampling time was taken from the sample identifier suffix, which encodes days 1, 3 and 5; we note that the deposited metadata contains no explicit timepoint field, so this mapping follows the cohort’s published sampling schedule rather than an annotated variable. Patients with at least two sampling timepoints and a recorded 28-day outcome entered the analysis (63 patients, 15 deaths). Exposures and the logistic model ladder were defined as in discovery, and odds ratios are reported per +1 standard deviation.

Sensitivity analyses in the validation cohort. Because a single validation estimate can be sensitive to analytical choices, we repeated the analysis under nine settings: the full sampling window; an early window restricted to days 1-3; a late window restricted to days 3-5; patients with complete day 1, 3 and 5 sampling; raw change rather than change per day; Firth penalised likelihood; an alternative module-scoring method based on across-sample gene-wise standardisation; and separately within the pneumonia and faecal-peritonitis strata. The early-window analysis was nominated, before any validation estimate was examined, as the setting most likely to recover a negative M3 association, because the discovery cohort with the largest event count sampled over a shorter interval.

Cross-modality triangulation (clinical analogues). Transcriptional module scores are not measured in routine care, so we additionally asked whether the biological direction of the two leading modules was reflected in routinely measured cell counts. We used MIMIC-IV v3.1, taking the first intensive-care admission of each adult meeting Sepsis-3 criteria (n=12,607; 28-day mortality 20.2%). For each patient we extracted white-cell differential measurements within 120 h of intensive-care admission and required at least two measurements separated by at least 12 h. The clinical analogue of M3 was the absolute lymphocyte count; the analogues of M1 were the neutrophil-to-lymphocyte ratio (computed within a single specimen), band forms and immature granulocytes.

Laboratory items were selected by explicit item identifier rather than by label matching, because the dictionary contains an atypical-lymphocyte item and a second absolute-lymphocyte item recorded in different units, either of which would be silently included by a text match. Change per 24 h was defined as for the transcriptomic analysis, and models were fitted unadjusted and adjusted for age, sex, first-day SOFA score and baseline value.

Interpretation of the cross-modality analysis. These analyses test whether the biological direction of a module is reflected in a different measurement modality; they do not validate the transcriptomic module scores themselves, and we describe them as convergent clinical evidence rather than as external validation of the signature.

### 2.8 Reproducibility, freezing, and provenance

The analysis is released as a frozen, end-to-end reproducible package. All analytical results are generated by a single committed pipeline script executed from a null-model reference through the full model ladder, meta-analysis, and figure generation, with a fixed random seed (42). After the two approved code corrections were committed (correct consumption of the precomputed OLS-slope z-score; restoration of the M0 null-model rows and the canonical 60-row × 27-column results schema), the pipeline was re-run end-to-end in a clean environment with no manual intervention and no aliasing of internal functions.

Every effect estimate, confidence interval, p value, q value and discrimination statistic in this manuscript is bound to an exact table row and column in a machine-readable evidence map (‘evidence_map.csv’); no value is recomputed for the manuscript. The frozen package includes the committed analysis code, the final frozen tables, the figures, a code diff, a reproducibility report, a machine-readable reproducibility verdict, a file manifest with SHA-256 checksums, an environment/package lock file, a data dictionary for all 27 result columns, a machine-readable evidence map binding each reported value to its source table, row and column, a script that verifies every binding against the frozen tables (‘verify_evidence_map.py’, S11 File), and a correction note documenting the bootstrap defect and its repair. Integrity of all frozen inputs was verified by SHA- 256 before and after manuscript preparation; the manifest accompanying this submission was regenerated against the final frozen tables. The original pre-fix artifacts are preserved unchanged in a clearly labelled archive and are explicitly not the reproducible analysis of record.

Core statistical results reproduced to machine precision across independent re-runs despite minor differences in installed package versions between the recorded environment metadata and the environment used for the final freeze; the true installed versions are recorded in the environment lock file, and the robustness of the inference to those version differences is noted.

Analyses were performed in Python (statsmodels, scikit-learn, pandas, numpy, scipy; lifelines and gseapy for supporting steps) as recorded in the environment lock file.

## 3. Results

### 3.1 Cohorts, feasibility, and descriptive dynamics

The three cohorts and their modeling feasibility are summarized in **Table 1** and **Table 2** (feasibility detail in S1 Table). Mortality modeling was feasible only in GSE95233 (51 patients; 17 non-survivors; EPV≈17) and GSE54514 (31 patients modeled; 7 events; EPV≈3.5); GSE57065 collected no mortality outcome and was reserved for early- dynamics analyses (26 complete H00/H24/H48 triplets). GSE95233 supported no multi-time-point slope (two samples per patient by design), so the OLS-slope exposure was confined to 29 GSE54514 patients (7 events).

**Table 2.** Primary per-cohort and cross-cohort effect estimates. Source: ‘per_cohort_effect_estimates.csv’ (per-cohort ORs, 95% CIs, p-values, and BH-FDR q-values for the model ladder) and ‘cross_cohort_meta_analysis.csv’ (pooled fixed-effect ORs, 95% CIs, p, q, and I²) for the change-per-24 h exposure, modules M1–M4. The table reports that no association reaches q<0.05 in any cohort or in meta-analysis; the minimum meta q is 0.075 (M3).

**(A) Per-cohort estimates — primary model (change per 24 h; OR per +1 SD).**
| Cohort | Module | n (events) | OR (per +1 SD change/24h) | 95% CI | p | q (BH-FDR) |
| --- | --- | --- | --- | --- | --- | --- |
| GSE95233 | M1 Emergency granulopoiesis | 51 (17) | 1.30 | 0.70–2.41 | 0.411 | 0.658 |
| GSE95233 | M2 Monocyte MHC-II suppression | 51 (17) | 0.59 | 0.31–1.12 | 0.108 | 0.217 |
| GSE95233 | M3 CD4/NK lymphocyte | 51 (17) | 0.56 | 0.30–1.05 | 0.071 | 0.190 |
| GSE95233 | M4 Interferon | 51 (17) | 1.16 | 0.64–2.11 | 0.626 | 0.696 |
| GSE54514 | M1 Emergency granulopoiesis | 31 (7) | 3.64 | 1.07–12.37 | 0.039 | 0.175 |
| GSE54514 | M2 Monocyte MHC-II suppression | 31 (7) | 2.01 | 0.75–5.36 | 0.164 | 0.262 |
| GSE54514 | M3 CD4/NK lymphocyte | 31 (7) | 0.44 | 0.16–1.24 | 0.120 | 0.240 |
| GSE54514 | M4 Interferon | 31 (7) | 1.20 | 0.51–2.84 | 0.675 | 0.675 |

**(B) Cross-cohort inverse-variance fixed-effect meta-analysis (change per 24 h; OR per +1 SD).**
| Module | Pooled OR (fixed-effect) | 95% CI | p | q (BH-FDR) | I<sup>2</sup> (%) |
| --- | --- | --- | --- | --- | --- |
| M1 Emergency granulopoiesis | 1.60 | 0.92–2.79 | 0.095 | 0.191 | 54 |
| M2 Monocyte MHC-II suppression | 0.85 | 0.50–1.46 | 0.564 | 0.564 | 76 |
| M3 CD4/NK lymphocyte | 0.53 | 0.31–0.90 | 0.019 | 0.075 | 0 |
| M4 Interferon | 1.17 | 0.72–1.92 | 0.522 | 0.564 | 0 |

Descriptively (Table 1), survivors and non-survivors differed most in the *change* of lymphocyte activity: in GSE95233, M3 (CD4/NK) rose in survivors (mean raw change +0.300) but fell in non-survivors (−0.284; Mann–Whitney p=0.029), whereas baseline M3 did not differ (p=0.757). In GSE54514, the raw change in M1 (immature neutrophils) was larger in non-survivors than survivors (+1.444 vs +0.082; p=0.012). These unadjusted descriptors motivated the trajectory modeling but are not corrected for multiplicity.

### 3.2 In discovery, no module-trajectory association survives FDR correction

Across the confirmatory family fixed before the analysis was run (M1–M4 × {baseline, change} within each mortality cohort; the plan had placed all five modules in it), no association reached the preregistered significance threshold after BH-FDR correction (all q>0.05; Table 2). Per-cohort, the smallest adjusted p- values were approximately q≈0.16–0.19 (M1-related terms). In the cross-cohort meta-analysis, the smallest adjusted p-value was q=0.075 (M3; Section 3.3). This is the central result: given 17 and 7 events, the study is powered to detect only large effects, and every signal below must be read as suggestive and hypothesis-generating rather than as a confirmed discovery.

### 3.3 Rising lymphocyte (M3) trajectory is the most consistent signal

The most consistent finding across cohorts was that a rising M3 (CD4/NK lymphocyte) trajectory was associated with lower mortality (Fig 1). In the inverse-variance fixed-effect meta-analysis of the change-per-24 h exposure, M3 gave a pooled OR of 0.53 (95% CI 0.31–0.90), p=0.019, with zero heterogeneity (I²=0%); because heterogeneity was null, the random-effects estimate was identical (OR 0.53, p=0.019). After BH-FDR correction this signal was not significant (q=0.075). The direction was concordant in both cohorts at the per-cohort level (GSE95233 change-model OR 0.56, p=0.071; GSE54514 change-model OR 0.44, p=0.120), consistent with the descriptive pattern of rising survivor lymphocyte activity (Section 3.1). This is a negative association between the change in M3 and mortality. Because the module has no uniquely determined functional orientation, it is described as such rather than as evidence of lymphocyte recovery or of a protective phenotype.

**Figure 1.**
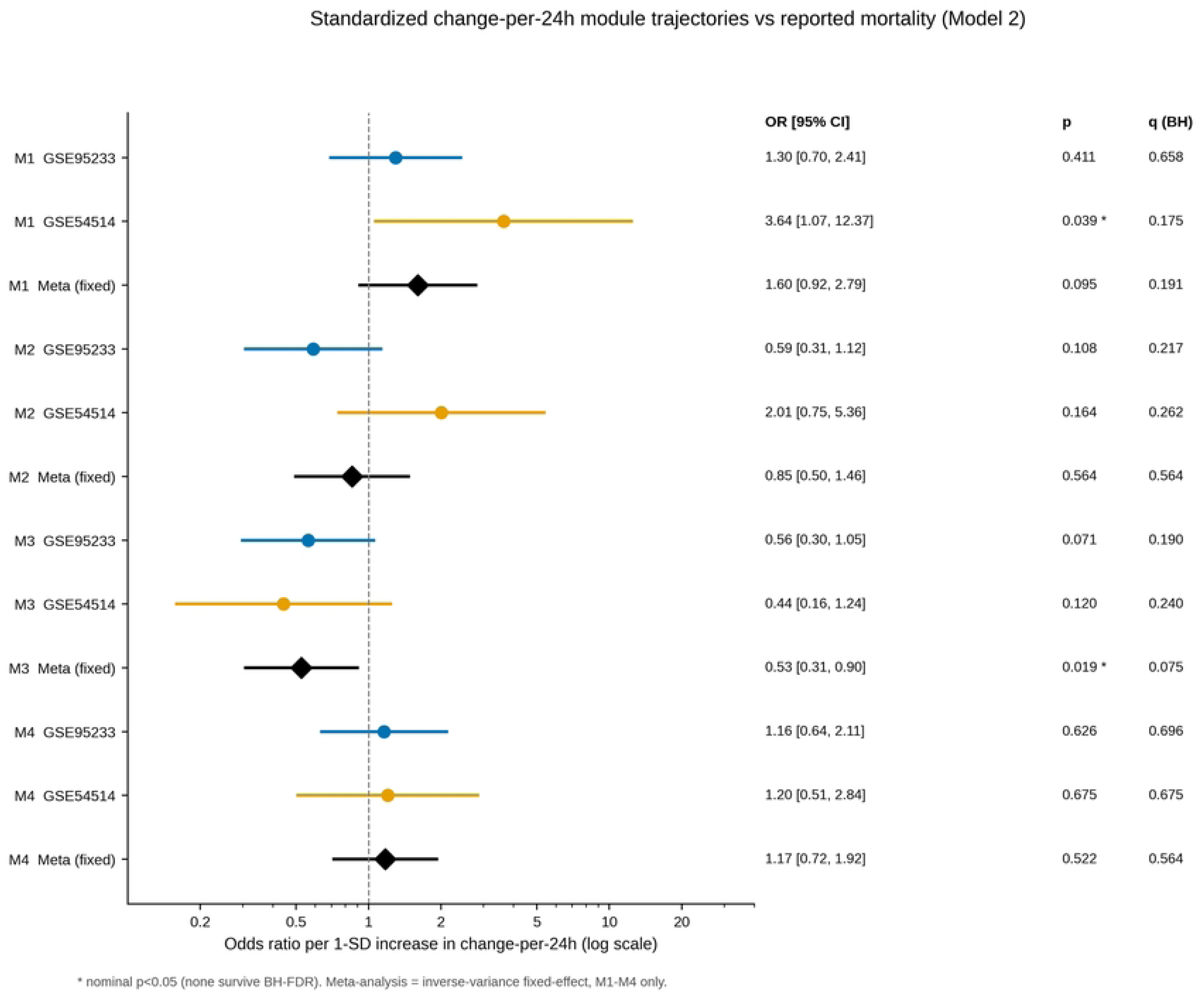
Cross-cohort meta-analysis of immune-module trajectories and mortality. Forest plot of inverse-variance fixed-effect pooled odds ratios (per +1 SD) for the change-per-24 h exposure of modules M1–M4 across the two mortality cohorts (GSE95233, GSE54514). Points are pooled ORs; whiskers are 95% confidence intervals; the vertical reference line marks OR = 1 (no association); the x-axis is on a log scale. The M3 (CD4/NK lymphocyte) module shows the only pooled OR whose confidence interval excludes 1 at the nominal level (OR 0.53, p=0.019), but it does not survive BH-FDR correction (q=0.075). Values are read directly from the frozen ‘cross_cohort_meta_analysis.csv’.

**Figure 2.**
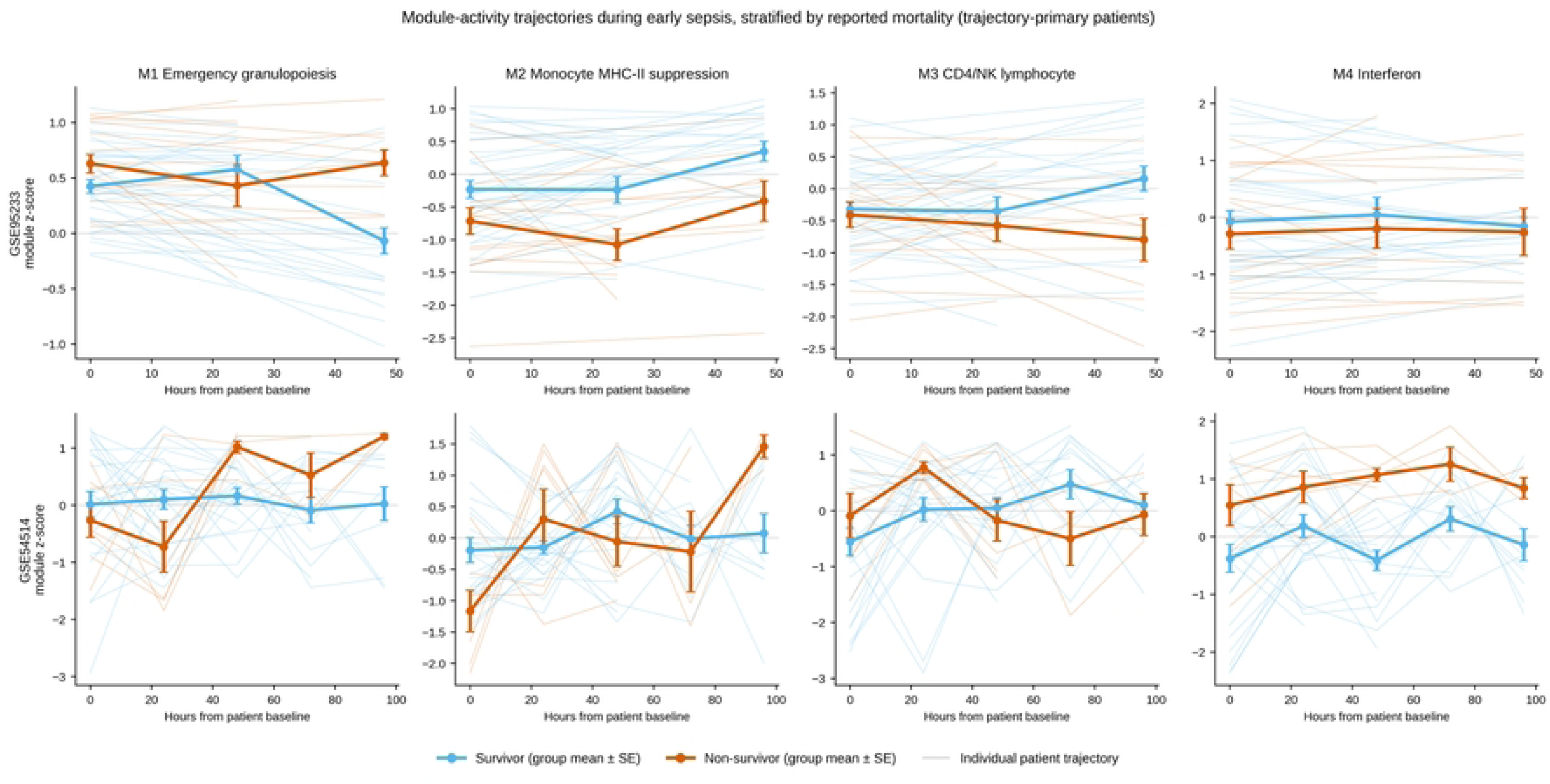
Immune-module trajectories by survival outcome. Module activity (z-scored) over the sampled early-sepsis interval in the mortality cohorts, stratified by survivor vs non-survivor. The panel for M3 illustrates the descriptive pattern of rising lymphocyte activity in survivors and falling activity in non-survivors that motivates the trajectory modeling. Trajectories depict frozen per-sample module scores; no smoothing or reanalysis was applied beyond the frozen pipeline.

### 3.4 Rising immature-neutrophil (M1) trajectory: nominal but heterogeneous and fragile

A rising M1 (emergency-granulopoiesis / immature-neutrophil) trajectory was nominally associated with higher mortality, but the pooled estimate was heterogeneous and imprecise (meta OR 1.60, 95% CI 0.92–2.79, p=0.095, q=0.191, I²=54%; Fig 1). The nominal signal was carried by GSE54514, where a rising M1 change was associated with higher mortality (change-model OR 3.64, 95% CI 1.07– 12.37, p=0.039, q=0.175). This M1 signal was influence-fragile in a cohort of 7 events (leave-one-out ORs ranged widely; S2 Table) and depended on the multi-day change rather than the earliest 24-h increment (it reversed under a two-time- point harmonization sensitivity analysis). It did, however, persist under APACHE-II adjustment in the flagged sensitivity model, indicating it carries information beyond baseline illness severity in that cohort. M2 (monocyte MHC-II suppression) pooled to a null and highly heterogeneous estimate (meta OR 0.85, p=0.564, q=0.564, I²=76%) driven by cohorts pointing in opposite directions, and M4 (interferon) was a consistent, well-behaved null (meta OR 1.17, p=0.522, q=0.564, I²=0%).

### 3.5 Trajectory shows nominal incremental association beyond baseline

Directly addressing the primary question, the model ladder indicated that within-patient change carried outcome information not captured by a single baseline snapshot in specific cohort–module instances, though imprecisely and without surviving FDR. In GSE54514, adding the M1 change to baseline improved model fit (LRT Model 3 vs Model 1 p=0.008), and the change-only model fit better than baseline by AIC (30.96 vs 36.72). In GSE95233, adding the M2 change improved fit over baseline (LRT Model 3 vs Model 1 p=0.007; Model 3 vs Model 2 p=0.004), with AIC decreasing from ∼65 to ∼59.5 (model-ladder detail in S3 Table). These improvements support the hypothesis that trajectory is informative, but they are demonstrated only in single cohort–module pairings and do not constitute confirmatory evidence.

### 3.6 Secondary slope analysis (GSE54514 only)

Because only GSE54514 had ≥3 serial samples, the OLS-slope exposure—the most direct test of *rate of change*—was a secondary, exploratory analysis in a single small cohort (n=29, 7 events) and is interpreted with caution. Directionally it agreed with the change-per-24 h results: a rising M1 slope was associated with higher mortality (Model-2 OR ≈ 10.89, 95% CI 1.43–82.87, p=0.021) and a rising M3 slope with lower mortality (Model-3 OR ≈ 0.18, 95% CI 0.034–0.977, p=0.047). The extremely wide confidence intervals, the small event count, proximity to separation, and the failure to survive FDR mean these slope estimates should be read as directionally supportive but quantitatively unreliable; the point ORs (and one near-degenerate interval in a fully adjusted slope model) should not be interpreted as effect-size estimates.

### 3.7 Early immune dynamics in a severity-only cohort (GSE57065)

In GSE57065 (no mortality outcome), early immune dynamics were directionally consistent with the mortality-cohort findings (Fig 3). In mixed-effects models over H00– H24–H48, M1 declined over the first 48 h in the pooled cohort (time slope −0.091, p=0.041). Illness severity modified the early lymphoid/monocyte trajectories: the time × SAPS-II interaction was significant for M2 (coefficient −0.289, p=0.004) and M3 (coefficient −0.347, p=0.012), and early-change (H00→H24) module shifts differed by SAPS-II severity for M2 (Mann–Whitney p=0.009, q=0.084) and M3 (p=0.017, q=0.084). Because this cohort has no outcome and n=26, these results are supportive plausibility signals only and none survives strict FDR.

**Figure 3.**
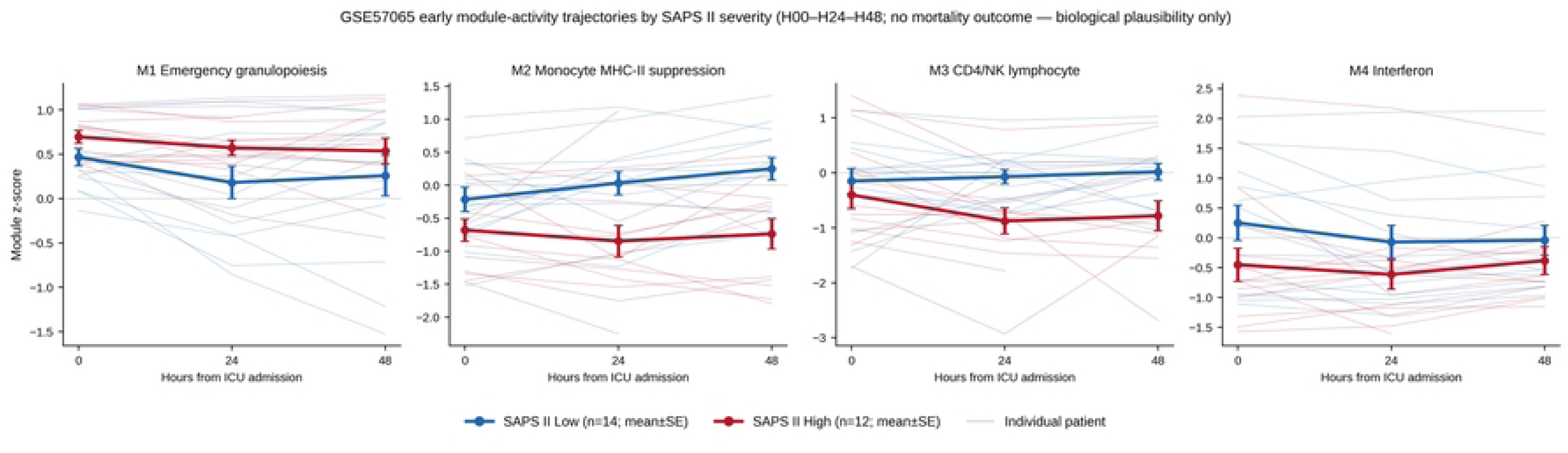
Early immune dynamics by illness severity in GSE57065. Module trajectories across H00, H24, and H48 in the severity-only cohort (no mortality outcome), stratified by binary SAPS-II severity (high vs low). M1 declines over the first 48 h (mixed-effects time slope p=0.041); the time × SAPS-II interaction is significant for M2 (p=0.004) and M3 (p=0.012). These are plausibility signals; none survives strict FDR (n=26).

### 3.8 Internal discrimination is modest and separation is rare

Apparent discrimination was modest and, as expected for in-sample estimates, optimistic: the best apparent AUC among the change-only models was 0.79 (GSE54514 M1; Fig 4), with most others near 0.5 to 0.68; across the whole ladder, including the models containing both baseline and change, apparent AUC reached 0.88 in the same cohort and module. After correction of the defect described in Section 2.6, optimism-corrected AUC was estimable for every model containing at least one exposure and, among the change-per-24 h models, ranged from 0.462 to 0.832 (including the exploratory OLS-slope models the range widens to 0.448 to 0.939); optimism was small, so corrected values lay close to apparent values, and several models were at or below chance. Separation was directly measured rather than assumed. Across the 10,000 bootstrap fits of the confirmatory change- exposure model (five modules x two cohorts x 1,000 resamples), separation was flagged by the fitting library in 9 (0.09%), all of them in the seven-event cohort, and every one still produced a finite linear predictor and a computable AUC; no fit failed to converge. The count refers to the separation diagnostic raised by the fitting library itself, which is a different and stricter test than the coefficient-magnitude rule used to trigger the penalised-likelihood fallback; under that coefficient rule the count is 10 of 10,000. In the original, non-resampled data no model was separated on any exposure. Calibration remains an in-sample recalibration (slope about 1.0, intercept about 0) and does not represent external calibration. These are internal discrimination estimates from the discovery cohorts; the external cohorts are used for association testing rather than for model transport.

**Figure 4.**
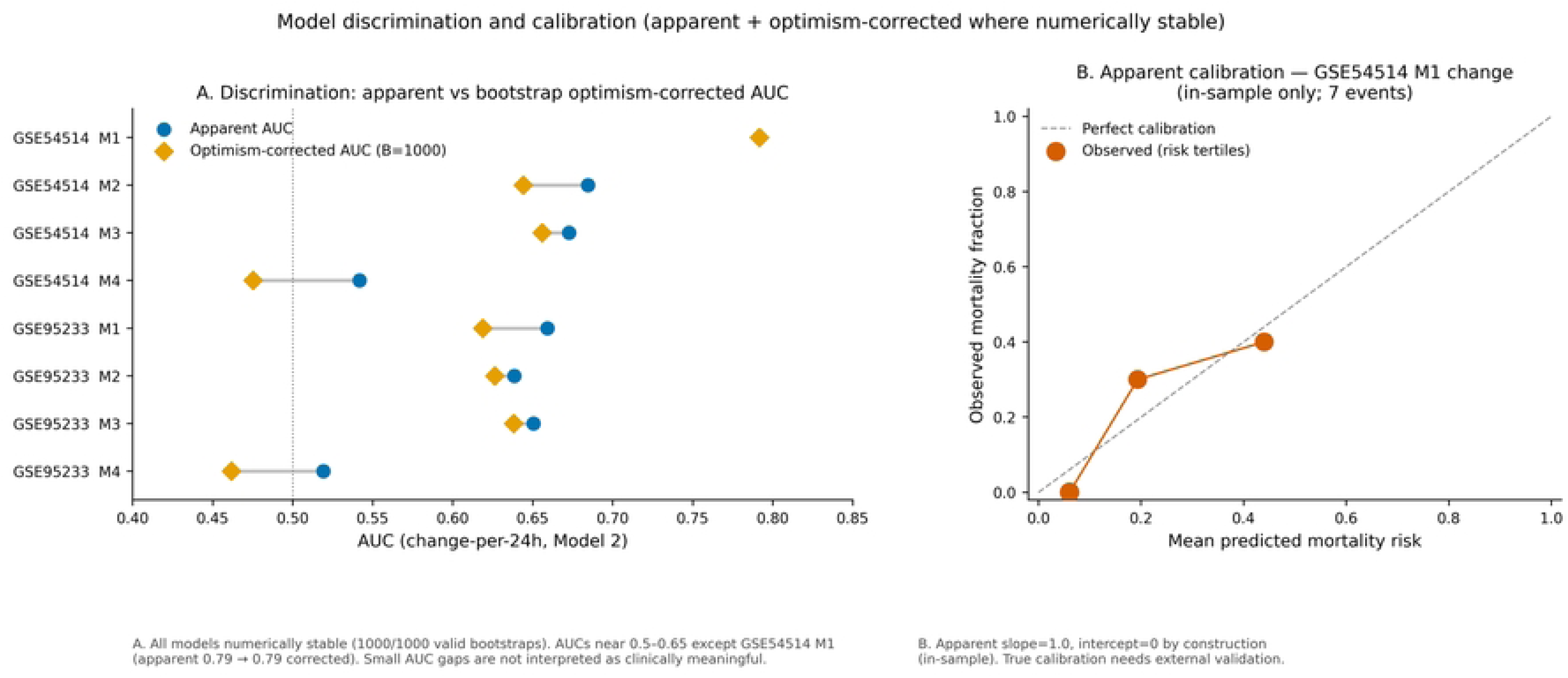
Apparent and optimism-corrected discrimination, and in-sample calibration. Apparent (in-sample) receiver-operating-characteristic performance, optimism-corrected AUC after bootstrap correction, and logistic recalibration for the discovery outcome models. Corrected discrimination was estimable for every model containing at least one exposure (M1–M3); the intercept-only M0 rung has no predictor and therefore no discrimination to correct. Several models were at or below chance. Calibration is shown as in-sample recalibration only (slope about 1.0, intercept about 0) and does not represent external calibration.

Restricting the analysis to a 48-hour landmark did not overturn either leading association. Among the patients whose last sample falls at 48 h, and who were therefore alive at the landmark, the pooled change-exposure odds ratio was 2.40 (0.98–5.89) for M1 and 0.30 (0.12–0.73) for M3, against 1.60 (0.92–2.79) and 0.53 (0.31–0.90) in the primary analysis; M2, M4 and M5 remained null (S2d Table). The landmark set is smaller (42 patients, 12 deaths, against 82 and 24) and the GSE54514 arm within it is very small, so these estimates are less precise rather than more reliable. What matters for the survivor-bias question is the direction: both associations moved slightly away from the null under the landmark, not toward it, which is not the pattern a pure immortal-time artefact would produce.

### 3.9 External transcriptomic validation: the M1 trajectory reproduces, the M3 trajectory does not

We applied the frozen modules and the identical analysis to the independent GAinS cohort (63 patients with serial sampling, 15 deaths; Fig 5). The two leading discovery signals behaved differently. The M1 (emergency-granulopoiesis) change association reproduced closely in both direction and magnitude: OR 1.72 (95% CI 0.92-3.20; p=0.088) in validation against 1.60 (0.92-2.79) in discovery. Pooling discovery and validation gave OR 1.65 (1.09-2.50; p=0.017) with no detectable heterogeneity (I-squared 0%). We emphasise that the validation estimate alone did not reach conventional significance; what reproduced was the direction and effect size, not a significance threshold.

**Figure 5.**
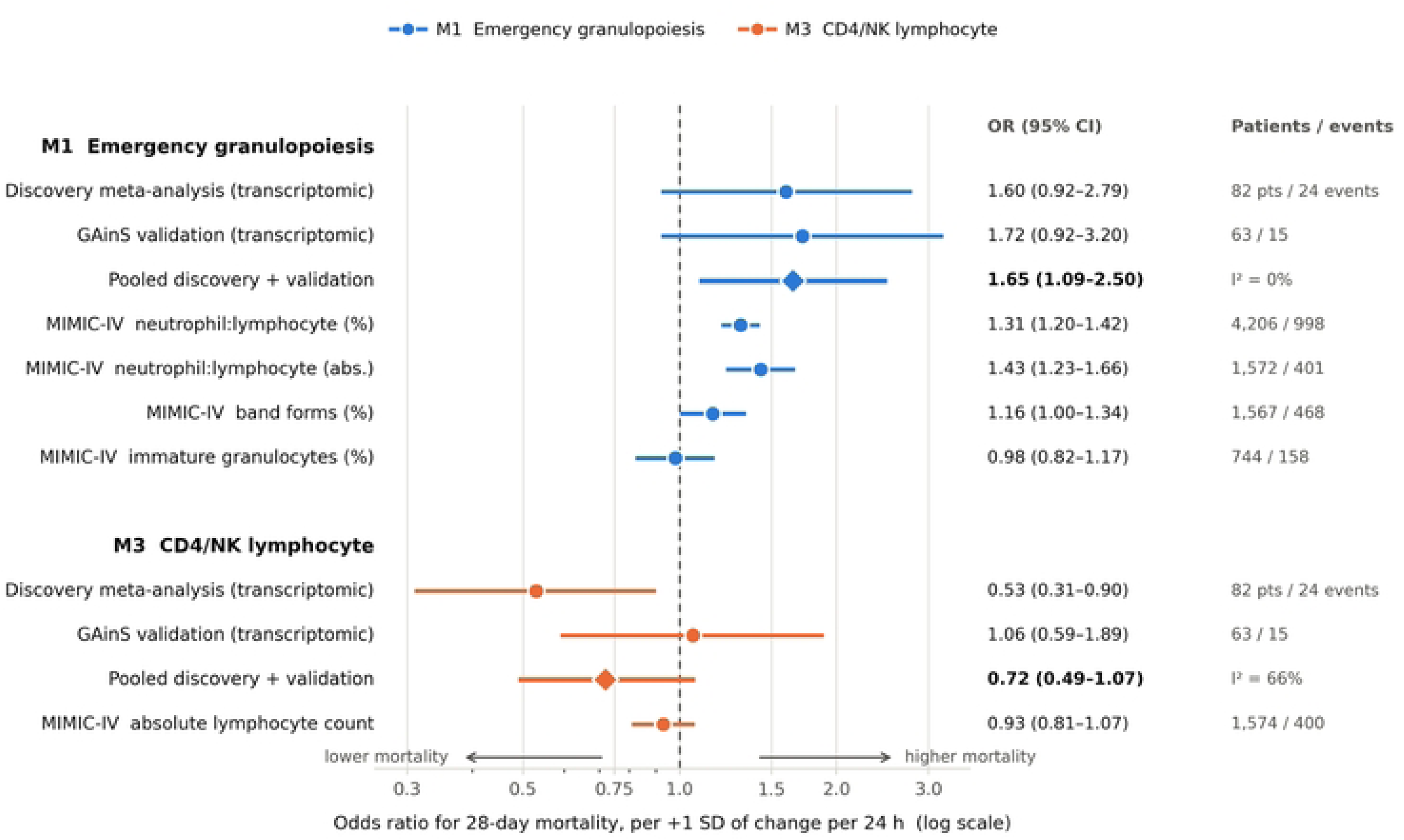
Discovery and independent validation of the two leading module trajectories. Odds ratios per +1 standard deviation of the change-per-24 h exposure for M1 (emergency granulopoiesis) and M3 (CD4/NK lymphocyte), shown for the discovery meta-analysis, the independent GAinS transcriptomic cohort, and the routine cell-count analogues in MIMIC-IV. Points are odds ratios, whiskers are 95% confidence intervals, the vertical reference line marks the null, and the x-axis is on a log scale. The M1 estimates are consistent in direction and magnitude across all sources; the M3 estimates are not.

The M3 (CD4/NK lymphocyte) change association did not reproduce. In validation the estimate was OR 1.06 (0.59-1.89; p=0.851), that is, essentially no association and in the opposite direction to discovery. Pooling discovery and validation gave OR 0.72 (0.49-1.07; p=0.11) with substantial heterogeneity (I-squared 66%), reflecting the disagreement between the two rather than a stable underlying effect. The remaining modules were consistent with discovery in being uninformative (M2 change OR 0.89; M4 change OR 0.78), while the exploratory M5 module moved in the same direction as M1 (OR 1.67).

The contrast was stable across the nine sensitivity settings, all of which were fixed before the estimates were examined. The M1 association was positive in all nine, with estimates confined to a narrow range (1.52 to 1.89): full window 1.72, early window 1.59, late window 1.57, complete day 1/3/5 sampling 1.52, raw change 1.60, Firth penalised likelihood 1.65, alternative scoring 1.58, pneumonia stratum 1.89 and faecal-peritonitis stratum 1.54. The M3 association was below 1 in only two of nine settings and ranged widely (0.44 to 2.17) without pattern. The setting nominated in advance as most favourable to a negative M3 association, the early day 1-3 window, gave OR 0.99, closer to the null than the main analysis. The non-reproduction of M3 is therefore not explained by dilution over a longer sampling interval.

### 3.10 Cross-modality triangulation: clinical analogues support M1 but not M3

In 12,607 adults meeting Sepsis-3 criteria in MIMIC-IV (28-day mortality 20.2%), we tested the routine cell-count analogues of the two leading modules using the same exposure definition and model structure (Table 3). The analogues of M1 supported the discovery direction. A rising neutrophil-to-lymphocyte ratio was associated with higher mortality (OR 1.31, 1.20-1.42, p=6.3e-10 using differential percentages, n=4,206 with 998 deaths; OR 1.43, 1.23-1.66, p=4.5e-06 using absolute counts, n=1,572 with 401 deaths), as was a rising band-form percentage (OR 1.16, 1.00- 1.34, p=0.048; OR 1.23, 1.05-1.43, p=0.010 after further adjustment for age, sex and first-day SOFA score). Not every analogue agreed: the immature-granulocyte percentage showed no association with mortality once baseline was accounted for (OR 0.98, 0.82-1.17, p=0.82), and we report this discordance rather than restricting attention to the supportive measures. We also note that the neutrophil-to-lymphocyte ratio trajectory was informative only after adjustment for its baseline value; the crude change was null (OR 1.05, p=0.14).

**Table 3.** Cross-modality triangulation in MIMIC-IV. Associations between the change per 24 h of routine white-cell measures and 28-day mortality among 12,607 adults meeting Sepsis-3 criteria. Odds ratios are per +1 standard deviation, from models containing baseline value and change, and from models additionally adjusted for age, sex and first-day SOFA score. The absolute lymphocyte count is the clinical analogue of module M3; the neutrophil-to-lymphocyte ratio, band forms and immature granulocytes are analogues of module M1. The baseline and change columns are the two terms of a single model; the adjusted column reports the change term from a model additionally containing age, sex and first-day SOFA score. Full model output, including baseline-only and change-only models, is provided in S6 Table.

| Routine white-cell analogue | Mod. | n | Deaths | Baseline OR (95% CI) | Change per 24 h OR (95% CI) | p | Adjusted change OR (95% CI) | p |
| --- | --- | --- | --- | --- | --- | --- | --- | --- |
| Neutrophil:lymphocyte ratio (differential %) | M1 | 4,206 | 998 | 1.50 (1.39–1.62) | 1.31 (1.20–1.42) | 6.3×10<sup>-10</sup> | 1.28 (1.17–1.39) | 2.9×10<sup>-8</sup> |
| Neutrophil:lymphocyte ratio (absolute counts) | M1 | 1,572 | 401 | 1.51 (1.32–1.72) | 1.43 (1.23–1.66) | 4.5×10<sup>-6</sup> | 1.40 (1.20–1.64) | 2.1×10<sup>-5</sup> |
| Band forms (%) | M1 | 1,567 | 468 | 1.01 (0.88–1.16) | 1.16 (1.00–1.34) | 0.048 | 1.23 (1.05–1.43) | 0.010 |
| Immature granulocytes (%) | M1 | 744 | 158 | 1.44 (1.22–1.70) | 0.98 (0.82–1.17) | 0.82 | 0.98 (0.82–1.18) | 0.85 |
| Absolute lymphocyte count (K/μL) | M3 | 1,574 | 400 | 0.93 (0.80–1.08) | 0.93 (0.81–1.07) | 0.31 | 0.97 (0.84–1.11) | 0.64 |

The clinical analogue of M3 gave a null result. A rising absolute lymphocyte count was not associated with mortality (OR 0.93, 0.81-1.07, p=0.31; n=1,574 with 400 deaths), and with 400 events the analysis had essentially complete power to detect a lymphocyte-count association of the magnitude estimated for M3 in discovery (OR 0.53) and 99.9% power to detect OR 0.70; the observed estimate differs from the discovery estimate at p=2.7e-15. This power statement applies to the lymphocyte count, not to the transcriptomic module. The absolute lymphocyte count measures circulating cell number; it carries no information about NK-cell function, cytotoxic-effector programmes or inhibitory-receptor expression, all of which contribute to M3, and it is therefore an incomplete analogue rather than a substitute measurement. The correct reading is that the absolute lymphocyte-count analysis did not provide cross-modality support for the M3 discovery association; it cannot exclude the module-level association, which is addressed by the transcriptomic validation in Section 3.9.

Taken together, three independent lines of evidence converge (Fig 5). The emergency-granulopoiesis trajectory reproduced in an independent transcriptomic cohort and was supported by two of its three clinical analogues in a cohort two orders of magnitude larger. The M3 CD4/NK trajectory failed to reproduce in the transcriptomic validation cohort, which used the identical frozen module and is therefore a direct external test, and failed in the sampling window most favourable to it. Its clinical analogue, the absolute lymphocyte count, did not provide cross-modality support at the discovery effect size; that count captures neither NK-cell function, cytotoxic programmes nor inhibitory-receptor expression, so it is an incomplete analogue of the module.

## 4. Discussion

*We asked whether the within-patient trajectory of defined immune-module activity during early sepsis is associated with mortality, and then subjected the two leading answers to independent testing. In discovery no association survived FDR correction; the two candidates were a rising CD4/NK lymphocyte-associated transcriptional signal associated with survival (pooled OR 0.53, q=0.075) and a rising emergency- granulopoiesis programme associated with death (pooled OR 1.60, q=0.191). Independent testing separated them, and in the opposite order to the one the discovery data suggested. The emergency-granulopoiesis association reproduced in an independent transcriptomic cohort (OR 1.72), in all nine sensitivity settings, and in the routine cell-count analogues of a cohort of 12,607 patients. The M3 association did not reproduce transcriptomically (OR 1.06), was null in the sampling window most favourable to it, and received no support from a well-powered clinical analogue*.

*The M3 finding deserves particular scrutiny, because it was the signal we would most readily have believed. It is directionally concordant with a substantial cellular literature: sepsis induces early depletion of CD4 T cells and NK cells, and both the depth of lymphopenia and the capacity to recover it are linked to survival [1, 9, 10, 11]. That coherence is precisely what made the discovery estimate persuasive despite a q value of 0.075. Yet it did not reproduce in the independent transcriptomic cohort, and it received no support from its clinical analogue. Three features of the discovery result look, in hindsight, like warning signs: it did not pass the prespecified FDR threshold; it rested on 24 events across two small cohorts; and its apparent consistency was driven by concordant direction rather than by precision. We think the most economical interpretation is that the discovery estimate was inflated by sampling variability in a small-event analysis, and that the biological literature on lymphocyte recovery, which concerns cell counts and function over longer horizons, does not translate into a detectable module-trajectory effect over the first days of sepsis at these sample sizes. A weaker effect than we estimated is not excluded by the transcriptomic data, and the clinical analogue cannot exclude a module-level effect of any size, because the absolute lymphocyte count does not measure the same construct; what it shows is that no lymphocyte-count association of the magnitude we reported for M3 is present in a well-powered cohort. The distinction is not academic. Lymphocyte-restoring therapy is an active line of investigation in sepsis, and a randomised trial has shown that interleukin-7 reverses the lymphopenia of septic shock [32]. If such an intervention is to be directed at the patients most likely to benefit, the markers used to select them need to survive exactly the kind of testing reported here*.

*The emergency-granulopoiesis finding, which we had described as fragile, proved to be the durable one. It fits an established framework in which emergency granulopoiesis and the accumulation of immature, immunosuppressive neutrophils mark the most severe septic state [14, 15]. What independent testing adds is that the trajectory, and not only the level, carries information: the association held after adjustment for baseline value and for illness severity, in a second transcriptomic cohort with a different case mix, and in two of three routine cell-count analogues. The discordant analogue, the immature- granulocyte percentage, is measured on a smaller subset and by an automated flag whose reproducibility across analysers is limited; we report the discordance rather than resolve it. The broader point is that a rising immature-neutrophil programme in the first days of sepsis is associated with worse outcome across measurement modalities, cohorts and case mixes, which is the pattern one expects of a real association rather than of a chance finding. The interferon module (M4) remained a clean null, and the monocyte MHC-II module (M2) remained uninterpretable across cohorts*.

Our question is adjacent to, but not answered by, the two closest recent studies. Serial whole-blood transcriptomic profiling with external mortality testing [1] establishes that a dynamic signature can transport across cohorts, but scores each sample on its own rather than asking whether within-patient change adds to a baseline value; its external panel includes E-MTAB-5273, the cohort used here for independent testing, so the two analyses are not fully independent of one another at the data level even though the modules and the estimand differ. Latent-class trajectory modelling of serial lymphocyte counts [33] tests a cell-count trajectory in far larger samples than are available to any transcriptomic study, and confirms it in MIMIC-IV, which is also the database used here for the clinical analogues. Its result — that lymphocyte-count trajectory phenotypes separate mortality — sits beside our null result for the absolute lymphocyte count and is not contradicted by it: a four-phenotype latent-class classification over four timepoints is a different exposure from a single standardized change per 24 hours, and the two can disagree without either being wrong. What neither study addresses, and what this one is built around, is the incremental question: whether early change carries outcome information beyond the baseline value of the same prespecified module, and whether that increment survives independent testing. Two methodological points follow. First, trajectory can be informative beyond baseline: in specific cohort-module pairings the baseline-plus-change model improved fit over baseline alone, and the same pattern appeared in the clinical analogues, where the neutrophil-to-lymphocyte trajectory was informative only once its baseline value was accounted for. Second, and more consequentially, the discovery signal that appeared most consistent was the one that failed. Had we reported the discovery analysis alone, the natural summary would have emphasised lymphocyte recovery, supported by a coherent literature and a q value just above threshold. The FDR threshold that we treated as an inconvenient technicality turned out to be the better guide. We would draw a narrow and practical conclusion from this: a trajectory association that does not clear its prespecified multiplicity threshold should be carried forward as a hypothesis to be tested, not as a finding to be interpreted, however biologically attractive it is.

### 4.1 Limitations

This study has important limitations. The discovery analysis remains small and exploratory, and the validation, while independent, has its own constraints.

1. **Few events and low EPV.** With 17 (GSE95233) and 7 (GSE54514) events, confirmatory power is minimal; no association survived FDR, and effect sizes— especially in GSE54514 and in the slope analysis—are imprecise with very wide confidence intervals.

2. Sparse events. With 17 and 7 events in the two discovery cohorts, effect estimates are imprecise and point estimates should be regarded as fragile. Separation, when directly measured during resampling, was rare (9 of 10,000 fits of the confirmatory change model) and never prevented estimation, but penalized-likelihood re-estimation remains a sensible check in this setting [29, 30]. An earlier version of this analysis reported the optimism-corrected discrimination as non-estimable because of separation; that attribution was a software defect and has been corrected, and the corrected values are reported throughout.

3. Association testing, not model transport. The external cohorts were used to test whether the trajectory associations reproduce, not to validate a prediction model. Discrimination and calibration remain internal to the discovery cohorts and are expected to be optimistic; we make no out-of-sample performance claim [19].

4. **Two-time-point change in the largest cohort.** GSE95233 supported only a change-per-24 h exposure, not a slope; the most direct rate-of-change test rested on a single small cohort (GSE54514, n=29, 7 events).

Survivor selection and informative sampling. Every trajectory analysis requires a patient to have contributed a second sample, which conditions the analysed set on surviving to the repeat draw, remaining in intensive care long enough for it, and having that measurement taken and pass quality control. The estimates are therefore open to immortal-time and survivor bias and to measurement-by-indication, which can change the magnitude and in principle the direction of a trajectory association rather than merely widening its confidence interval. The written plan proposed a landmark framing at 48–72 h as a mitigation. None of the discovery cohorts records time to death, only survival status at the end of follow-up, so patients who died before a landmark cannot be identified directly; they can, however, be excluded by construction, because a patient with a sample drawn at 48 h was alive at 48 h. The 48-hour landmark analysis reported in Section 3.8 and S2d Table does this, and neither leading association reverses or disappears under it. That result bounds the immortal-time component of the bias. It does not remove measurement-by-indication: the landmark set is conditioned on having been re- sampled, not merely on being alive, and patients who were re-sampled may still differ systematically from those who were not. A 72-hour landmark is not estimable, because no GSE95233 patient has a sample beyond 48 h. The comparison of included with excluded patients (S2c Table) bounds a further part of the problem but does not remove it. The limitation applies to the discovery and transcriptomic testing stages alike; it is one reason the routine cell-count analogues, where serial measurement is ordinary clinical practice rather than a study protocol, add information even though they measure a different construct.

5. **Cross-platform, cross-cohort heterogeneity.** The cohorts differ in microarray platform, sampling schedule, severity mix, and outcome ascertainment; observed heterogeneity (e.g., M1 I²=54%, M2 I²=76%) limits pooling, and the exact mortality horizon for GSE54514 could not be resolved (coding is binary and horizon-independent, so no estimate changes, but comparability across cohorts is imperfect).

6. Single-sample module scoring. Module activity was quantified by within-sample rank-percentile scoring, with single-sample enrichment [23, 24] as the alternative-scoring sensitivity analysis; both are single-sample approaches, which are sensitive to platform, normalization, and gene-set composition and may not reproduce group-level separation seen with pairwise methods, as evaluated in method comparisons including two preprints [25, 26] (preprints; not peer-reviewed); module definitions were frozen a priori to avoid data-driven optimism, but residual scoring dependence remains.

7. **Severity-only third cohort.** GSE57065 has no mortality outcome; its dynamics are plausibility support, not outcome evidence, and its severity associations (n=26) do not survive strict FDR.

8. Exploratory M5, its construct change, and secondary analyses. M5 was designated exploratory and excluded from confirmatory testing and from pooling. That restriction is not present in the preregistered plan and was taken by the analysts at the frozen stage. An earlier internal description of the pipeline attributed the exclusion to the module being available on one microarray platform only; that attribution is incorrect, since twelve of its thirteen reference genes map to both platforms and the module was scored and fitted in every cohort, and it has been corrected here and in the supporting documentation. Separately, the plan defined M5 as a contrast between persistent innate inflammation and a recovery or adaptive axis, whereas the implemented score contains only same-direction innate-inflammatory genes; the module therefore measures innate-inflammatory transcriptional activity and must not be read as inflammation-versus-recovery. Both departures are itemised in the deviation log. All M5 estimates were null in both mortality cohorts and are reported in full in the supporting information, so neither departure affects any reported conclusion; including M5 in the confirmatory family would have made the multiplicity correction more stringent, not less, and no association survived it as reported. The slope analysis was prespecified as the primary exposure and appears here as a secondary analysis, because only one cohort has the three or more serial samples it requires (S8 Appendix, item 1.1); the APACHE-II-adjusted models were prespecified as a sensitivity analysis. Neither is confirmatory.

Unadjusted models and deviations from the preregistered plan. The reported model ladder contains no demographic covariates: with 17 and 7 events, the event-per-variable budget did not permit adding age and sex alongside two module terms, and the preregistered plan had specified age plus one module term as the default adjusted model. All reported estimates are therefore unadjusted for demography and answer whether the trajectory is associated with mortality, not whether it is associated independently of age and sex. The plan had also named the change in discrimination between the baseline-plus-change and baseline-only models as the primary readout; that comparison is reported as a likelihood-ratio test and a model-ladder table rather than as a bootstrap-interval difference in discrimination, and the effect estimates are foregrounded instead. These and all other differences from the plan, including the prespecified sensitivity analyses that were not executed and the two validation stages that were added after the discovery analysis, are itemised in the deviation log provided as supporting information.

9. **No causal or clinical-utility inference.** All associations are observational; none supports causal interpretation, and none has been evaluated for clinical decision-making.

10. Limitations of the transcriptomic validation. The GAinS validation cohort contributed 63 patients and 15 deaths, comparable to the largest discovery cohort rather than substantially larger, so the validation estimate for M1 is itself imprecise and did not reach conventional significance on its own. The deposited metadata contains no explicit timepoint field, and we inferred days 1, 3 and 5 from the sample identifiers in line with the cohort’s published sampling schedule; an error in this mapping would rescale the change-per-day exposure, although it would not reverse its direction. The case mix (community-acquired pneumonia and faecal peritonitis) differs from the discovery cohorts, which is a strength for generalisability and a limitation for exact comparability.

11. Limitations of the clinical-analogue triangulation. Cell counts are not module scores. A neutrophil-to-lymphocyte ratio reflects circulating cell proportions, whereas M1 reflects a transcriptional programme of granulopoiesis; agreement in direction supports the biology but does not validate the signature, and disagreement would not by itself refute it. The MIMIC-IV analyses are retrospective, single-centre in origin, and subject to measurement-by-indication, since patients who have differential counts repeated are not a random subset. One of three M1 analogues was null. The M1 analogues are also subject to the same cellular-composition ambiguity that applies to M3: a neutrophil-to-lymphocyte ratio or a band count reflects the proportion and maturity of circulating cells and cannot by itself confirm that an emergency-granulopoiesis transcriptional programme is what is changing. The lymphocyte analogue was well powered against the discovery effect size but cannot exclude a small effect.

Adults only. Every cohort analysed here, in both the discovery and the validation stages, comprises adults. Whole-blood transcriptomic heterogeneity has also been described in paediatric septic shock, where distinct subclasses carry different outcomes [31], and the immune trajectories of early sepsis need not behave the same way in children. Whether the associations reported here extend below adulthood is untested.

12. No causal or clinical-utility inference. All associations are observational. Nothing here establishes that emergency granulopoiesis causes death rather than marking severity, and no measure reported here has been evaluated for clinical decision-making.

### 4.2 Conclusions

*Within-patient immune-module trajectories during early sepsis were examined in three discovery cohorts and then tested independently. A rising emergency-granulopoiesis programme was associated with higher mortality, and this association was directionally reproduced in an independent transcriptomic cohort, where it was of similar size but did not reach conventional significance on its own (OR 1.72, 0.92 to 3.20, p=0.088), was positive across all nine sensitivity settings, and was supported by the routine cell-count analogues drawn from a database of 12,607 patients, in which between 744 and 4,206 contributed the serial measurements each analogue required. A rising CD4/NK*

*lymphocyte-associated transcriptional signal, which was the more consistent-looking result in discovery and the more biologically expected one, did not reproduce, and its clinical analogue, the absolute lymphocyte count, provided no cross-modality support, although that count measures a different construct and cannot by itself exclude a module-level association. Neither discovery association had cleared the prespecified FDR threshold, and independent testing showed that this threshold, rather than biological plausibility, was the better guide to which signal would survive. The findings support emergency granulopoiesis trajectory as a candidate marker worth prospective evaluation and argue against the M3 CD4/NK trajectory at the effect size we originally estimated. This study is observational and establishes neither causality nor clinical utility*.

## Supporting information

The following are provided as Supporting Information. The effect estimates and meta-analysis are read directly from the frozen analysis package. The feasibility, sensitivity, severity and stability tables are regenerated from the frozen module-score inputs by a committed script, because the released pipeline executed only its first four stages and these tables were not deposited with it; the regenerated values reproduce the reported ones. The specification search for the severity-only cohort (S4c Table) and the separation-counting variants (S5c Table) are new analyses performed to document choices that the original run had not recorded.

- S1 Appendix. Supplementary methods. Full module gene membership (‘modules_frozen.json’), cohort provenance and outcome-metadata verification, exposure standardization, the complete model/meta/FDR specification, the GAinS scoring and timepoint-inference procedure, and the MIMIC-IV extraction specification; data dictionary for all 27 result columns (‘data_dictionary_per_cohort_effect_estimates.csv’); correction note for the bootstrap defect (‘CORRECTION_NOTE.md’).
- S1 Table. Event and model feasibility. Per cohort and module: total patients, patients with two or more serial samples, patients meeting the trajectory- primary criterion, modelled patients, 28-day deaths, events per variable, median follow-up, and whether the change model was fitted (‘S1_event_model_feasibility.csv’).
- S2 Table. Discovery sensitivity analyses. Odds ratios for the change exposure under six settings — primary change per 24 h, raw change, forced Firth penalised likelihood, alternative single-sample scoring (ssGSEA), APACHE II adjustment in GSE54514, and harmonisation to the first two timepoints — together with leave-one-out influence and an informative-dropout comparison of included versus excluded patients (‘S2_sensitivity_analysis_matrix.csv’, ‘S2b_leave_one_out_influence.csv’, ‘S2c_informative_dropout.csv’).

S2b Table. Leave-one-out influence. For every cohort and module, the change- exposure odds ratio refitted with each patient omitted in turn, with the full-sample estimate, the minimum, median and maximum leave-one-out estimates, the fold range, the most influential patient and whether removing any single patient moves the estimate across the null (‘S2b_leave_one_out_influence.csv’).

S2c Table. Informative dropout. Patients meeting the trajectory-primary criterion compared with those excluded, on baseline module score and on mortality, with Mann-Whitney and Fisher tests (‘S2c_informative_dropout.csv’).

S2d Table. Landmark sensitivity analysis at 48 hours. The change-exposure odds ratio for every cohort and module, refitted among patients whose last frozen sample falls at 48 hours and who were therefore alive at the landmark, with exposures re-standardised within that set, reported beside the corresponding primary estimate and pooled by inverse-variance fixed effect (‘S2d_landmark_48h_sensitivity.csv’).

S3 Table. Full model ladder. Model 0 to Model 3 for every cohort, module and exposure, with AIC and the change in AIC against the intercept-only model, likelihood-ratio tests, apparent and optimism-corrected discrimination, odds ratios with confidence intervals, and Benjamini-Hochberg q values (‘S3_model_ladder_M0_M3.csv’, and the complete per-cohort estimate table ‘per_cohort_effect_estimates.csv’).

S4 Table. GSE57065 severity analyses. Baseline module scores by SAPS II stratum with Mann-Whitney tests, and linear mixed-effects trajectory models (module score on time, severity stratum and their interaction, with a random intercept per patient) (‘S4_GSE57065_severity_associations.csv’, ‘S4b_GSE57065_mixedeffects_trajectories.csv’).

S4b Table. GSE57065 mixed-effects trajectories. Coefficients, standard errors and p-values from two linear mixed-effects models per module: a pooled time-trend model containing time alone, and a model containing time, SAPS-II stratum and their interaction. Both use complete H00/H24/H48 triplets, a random intercept per patient and maximum-likelihood estimation (‘S4b_GSE57065_mixedeffects_trajectories.csv’).

S4c Table. GSE57065 specification search. Eight candidate mixed-effects specifications (all patients versus complete triplets, random intercept versus random intercept and slope, restricted versus full maximum likelihood) with their distance from the reported coefficients, documenting how the specification in S4b Table was identified (‘S4c_GSE57065_specification_search.csv’).

S5 Table. Bootstrap and calibration stability. Apparent and optimism-corrected AUC, the optimism estimate, valid-replicate counts, and in-sample recalibration slope and intercept for every fitted model; and the directly measured incidence of complete or quasi-complete separation across bootstrap resampling (‘S5_stability_bootstrap_calibration.csv’, ‘S5b_separation_incidence.csv’).

S5b Table. Separation incidence by model. The number of bootstrap resamples in which the fitting library raised a separation diagnostic, per cohort and module, together with the number of degenerate resamples in which all resampled patients shared one outcome (‘S5b_separation_incidence.csv’).

S5c Table. Separation counting variants. The same count under two criteria (the fitting library’s own separation diagnostic and the coefficient-magnitude rule used to trigger the penalised-likelihood fallback) crossed with two resampling-stream conventions, documenting the sensitivity of the reported incidence to these choices (‘S5c_separation_counting_variants.csv’).

S6 Table. Cross-modality triangulation model ladder (MIMIC-IV). Complete model output for every routine white-cell analogue in Table 3: baseline-only, change- only, baseline-plus-change, and the model additionally adjusted for age, sex and first-day SOFA score, with odds ratios, 95% confidence intervals, p-values, patient counts and event counts (‘S6_mimic_model_ladder.csv’). The MIMIC-IV patient- level table cannot be redistributed under the data use agreement; the extraction query in the code archive regenerates it for any credentialed user.

S7 Table. External transcriptomic validation sensitivity analyses (GAinS). The nine settings reported in Section 3.9, each fixed before the validation estimates were examined — full window, early window, late window, complete day 1/3/5 sampling, raw change, Firth penalised likelihood, alternative scoring, pneumonia stratum and faecal-peritonitis stratum — with the odds ratio and 95% confidence interval for M1 and M3 in each (‘S7_gains_sensitivity.csv’).

S7b Table. External transcriptomic validation, full model results (GAinS). Odds ratios, 95% confidence intervals, p-values and Benjamini-Hochberg q-values for the baseline, change and baseline-plus-change models of all five modules in the validation cohort, with patient and event counts (‘S7b_gains_module_results.csv’).

S8 Appendix. Deviations from the preregistered analysis plan. An itemised comparison of the analysis plan with the analysis reported here, covering changes forced by cohort structure, changes chosen by the authors, prespecified analyses that were not executed, and the two validation stages added after the discovery analysis was complete.

S9 File. Preregistered analysis plan. The written analysis plan in full, as finalised before any outcome model was run. Two dated resolution notes concerning the GSE54514 outcome horizon were added before manuscript preparation and are marked within the document.

S10 File. Frozen module definitions. The machine-readable file that defines the five immune modules (‘modules_frozen.json’), frozen before any outcome model was run: for each module the reference gene list, the genes removed as ribosomal, mitochondrial or haemoglobin transcripts, platform availability on GPL570 and GPL6947, the final usable gene list used for scoring, and the derivation note. All module scores in this manuscript are computed from the final usable gene lists in this file.

S11 File. Evidence-map verification script. A Python script (‘verify_evidence_map.py’) that reads the evidence map and the frozen result tables supplied with this submission, resolves every binding to its table row and column, compares the resolved value with the value as reported in the manuscript, and exits non-zero if any binding fails to resolve or to match. Run as ‘python verify_evidence_map.py <directory containing the supporting tables>’.

S12 File. Landmark sensitivity script. The script that produces S2d Table (‘landmark_sensitivity.py’), together with the frozen patient-level analysis dataset it reads (‘patient_level_analysis_dataset.csv’: one row per cohort, patient and module, carrying the frozen baseline score, the change per 24 h, the sampling times, the trajectory-primary flag and the outcome), so that the analysis can be rerun from the submitted files alone. It fits the logistic models directly and, run in its primary setting, reproduces every per-cohort change-exposure odds ratio in ‘per_cohort_effect_estimates.csv’ to within 1e-9, which is the check that it fits the same models as the frozen pipeline.

S13 File. STROBE checklist. The STROBE checklist for cohort studies, completed for this manuscript, giving for each of the 22 items the manuscript section, display item or supporting-information file in which it is reported, and marking explicitly the items that do not apply to a secondary analysis of existing cohorts.

- ·**Repository-deposited data (cited from the Data Availability statement, not uploaded as journal supporting information): the frozen per-patient exposure** tables and per-sample module-score tables for all three discovery cohorts, the GAinS module-score table, and the alternative single-sample (ssGSEA) scoring table (‘exposures_alt_ssgsea.csv’). These derived data are archived in Zenodo at 10.5281/zenodo.21679713.

No supplementary figures are provided; all display items are the five main figures and three main tables.

## 8. Declarations

### Data availability

The discovery transcriptomic datasets are available from the Gene Expression Omnibus under accessions GSE95233, GSE54514 and GSE57065. The external transcriptomic validation cohort is available from ArrayExpress/BioStudies under accession E-MTAB-5273. The cross-modality triangulation used MIMIC-IV v3.1, which is available to credentialed users from PhysioNet (https://physionet.org/content/mimiciv/) under its data use agreement; access requires completion of the PhysioNet credentialing process and human-subjects research training, the same route the authors used, and the authors hold no special access. The complete extraction query is provided in the code archive so that the cohort can be regenerated exactly by any credentialed user. The curated derived datasets supporting the reported analyses are archived in Zenodo at 10.5281/zenodo.21679713 (corrected release; the earlier record 10.5281/zenodo.21496589 contains the pre-correction artifacts and is superseded).

## Code availability

The analysis code, reproducibility documentation, computational environment specifications, and associated derived-data files are available through GitHub at https://github.com/sulei1987-sudo/sepsis-immune-trajectory-analysis and archived in Zenodo at 10.5281/zenodo.21679713. The pre-correction version of the pipeline is archived unchanged at 10.5281/zenodo.21496589 and is explicitly not the analysis of record.

## Reproducibility statement

Every effect estimate, confidence interval, p value, q value and discrimination statistic reported here is bound to its source table, row and column in a machine-readable evidence map (‘evidence_map.csv’); the map is regenerated from the frozen tables by a committed script (‘verify_evidence_map.py’, provided as S11 File) that resolves each binding to its table row and column and confirms the value against the frozen table, so it stays in step with the paper. Each binding is located in the manuscript by matching the value together with its cohort, module or measure rather than by the numeric string alone, so a number that occurs in more than one place cannot be bound to the wrong sentence; candidate values that cannot be located under that rule are reported as not present in the text rather than given a guessed location. No value was recomputed for the manuscript. The integrity of all frozen inputs was verified by SHA-256 checksums (‘SHA256SUMS_outputs.csv’), regenerated against the final frozen tables after manuscript preparation; the manifest covers every result table submitted with this manuscript. A software defect in the bootstrap optimism-correction helper, which had caused the optimism-corrected AUC to be reported as non-estimable, was identified on audit and corrected; the correction note (‘CORRECTION_NOTE.md’) and the output checksums (‘SHA256SUMS_outputs.csv’) are provided with this submission, and ‘modules_frozen.json’ (frozen module gene membership) is provided as S10 File, and every other file named in this manuscript is in the public code archive and the Zenodo derived-data deposit; all discrimination values reported here come from the corrected computation, and the defect is documented in full in a companion methodological report and in the code archive. The original pre-fix artifacts are archived unchanged and are explicitly not the analysis of record.

## Author contributions

Lei Su: Conceptualization, Methodology, Data curation, Formal analysis, Visualization, Writing – original draft, and Writing – review and editing. Lin Zhang: Interpretation of results and Writing – review and editing. Wencai Huang: Interpretation of results and Writing – review and editing. Chunmei Gui: Supervision, Funding acquisition, Interpretation of results, and Writing – review and editing. Fang Gong: Supervision, Interpretation of results, and Writing – review and editing. All authors approved the final version of the manuscript and agree to be accountable for the work.

## Ethics

This study involved secondary analysis of existing, de-identified human data and generated no new patient data. The discovery transcriptomic datasets were obtained from the Gene Expression Omnibus (GSE95233, GSE54514 and GSE57065) and the external transcriptomic validation cohort from ArrayExpress/BioStudies (E-MTAB-5273); all are publicly available and de-identified, and the original studies that generated them were conducted with the ethical approvals and participant consent reported in their source publications. The cross-modality analyses used MIMIC-IV v3.1, a de-identified critical-care database distributed by PhysioNet to users who have completed its credentialing process and human-subjects research training and have signed its data use agreement; the authors accessed it by that route and hold no special access. MIMIC-IV was approved for public release with a waiver of informed consent by the institutional review board of the Beth Israel Deaconess Medical Center. Ethical approval was not required for the present study, because it involved secondary analysis of de-identified data with no direct interaction with human participants, no collection of human samples, and no access to identifiable human information.

## Data Availability

No new patient data were generated by this study all analyses used existing, publicly available or credentialed-access data sources. The discovery transcriptomic datasets are available from the NCBI Gene Expression Omnibus under accessions GSE95233 (https://www.ncbi.nlm.nih.gov/geo/query/acc.cgi?acc=GSE95233), GSE54514 (https://www.ncbi.nlm.nih.gov/geo/query/acc.cgi?acc=GSE54514) and GSE57065 (https://www.ncbi.nlm.nih.gov/geo/query/acc.cgi?acc=GSE57065). The external transcriptomic validation cohort is available from ArrayExpress/BioStudies under accession E-MTAB-5273 (https://www.ebi.ac.uk/biostudies/arrayexpress/studies/E-MTAB-5273). The cross-modality analyses used MIMIC-IV v3.1, available to credentialed users from PhysioNet (https://physionet.org/content/mimiciv/) under its data use agreement. Access requires completion of the PhysioNet credentialing process and human-subjects research training this is the route the authors used and the authors hold no special or privileged access. The complete extraction query is deposited with the analysis code so that the cohort can be regenerated exactly by any credentialed user. The curated derived datasets underlying every reported result - the frozen per-patient exposure tables and per-sample module-score tables for all three discovery cohorts, the validation-cohort module-score table, and the alternative-scoring table - are archived in Zenodo at 10.5281/zenodo.21679713. The analysis code, the frozen result tables, the computational environment specification, and a machine-readable evidence map binding every reported value to its source table row and column are available at https://github.com/sulei1987-sudo/sepsis-immune-trajectory-analysis and archived in the same Zenodo record.

https://doi.org/10.5281/zenodo.21679713

https://physionet.org/content/mimiciv/

https://www.ebi.ac.uk/biostudies/arrayexpress/studies/E-MTAB-5273

https://www.ncbi.nlm.nih.gov/geo/query/acc.cgi?acc=GSE57065

https://www.ncbi.nlm.nih.gov/geo/query/acc.cgi?acc=GSE54514

https://www.ncbi.nlm.nih.gov/geo/query/acc.cgi?acc=GSE95233

Full bibliographic records for all 33 cited sources, numbered sequentially by first appearance in the text. Author lists, article titles, journals, and DOIs are provided as captured from the source records. Volume, issue, and page or article numbers were backfilled from Crossref (primary) and PubMed/NCBI (secondary cross-check) and validated by DOI, title, journal, author, and year matching; PubMed (PMID) and PubMed Central (PMCID) identifiers are included where available. References 25 and 26 are preprints (not peer-reviewed) with no peer-reviewed journal version and therefore have no volume, issue, or pages. DOIs resolve to the definitive record of each article.

## References

1. Hwang J, Cha JH, Ahn S, Park JH, Kim S, Jung M, Choi Y, Seok H, Choi WS, Nam MH, Park DW. Longitudinal blood transcriptome profiling reveals immune dynamics in sepsis. BMC infectious diseases. 2026;26(1):1229. doi:10.1186/s12879-026-13416-1. https://pubmed.ncbi.nlm.nih.gov/42106604/ PMID:42106604; PMC13335145.

2. Cabrera-Perez J, Condotta SA, Badovinac VP, Griffith TS. Impact of sepsis on CD4 T cell immunity. Journal of leukocyte biology. 2014;96(5):767–777. doi:10.1189/jlb.5MR0114-067R. https://pmc.ncbi.nlm.nih.gov/articles/PMC4197564/ PMID:24791959; PMC4197564.

3. E. Cano-Gamez, K. Burnham, Cyndi Goh, et al.. An immune dysfunction score for stratification of patients with acute infection based on whole blood gene expression. Science translational medicine. 2022;14(669):eabq4433. doi:10.1126/scitranslmed.abq4433. 10.1126/scitranslmed.abq4433 PMID:36322631; PMC7613832.

4. Sweeney TE, Azad TD, Donato M, Haynes WA, Perumal TM, Henao R, Bermejo- Martin JF, Almansa R, Tamayo E, Howrylak JA, Choi A, Parnell GP, Tang B, Nichols M, Woods CW, Ginsburg GS, Kingsmore SF, Omberg L, Mangravite LM, Wong HR, Tsalik EL, Langley RJ, Khatri P. Unsupervised Analysis of Transcriptomics in Bacterial Sepsis Across Multiple Datasets Reveals Three Robust Clusters. Crit Care Med. 2018;46(6):915–925. doi:10.1097/CCM.0000000000003084. 10.1097/CCM.0000000000003084 PMID:29537985; PMC5953807.

5. G. Parnell, B. Tang, M. Nalos, et al.. Identifying Key Regulatory Genes in the Whole Blood of Septic Patients to Monitor Underlying Immune Dysfunctions. Shock. 2013;40(3):166–174. doi:10.1097/shk.0b013e31829ee604. 10.1097/shk.0b013e31829ee604 PMID:23807251.

6. Shayantan Banerjee, Akram Mohammed, H. Wong, et al.. Machine Learning Identifies Complicated Sepsis Course and Subsequent Mortality Based on 20 Genes in Peripheral Blood Immune Cells at 24 H Post-ICU Admission. Frontiers in Immunology. 2021;12:592303. doi:10.3389/fimmu.2021.592303. 10.3389/fimmu.2021.5923033 PMID:33692779; PMC7937924.

7. Anne M. Drewry, Enyo A. Ablordeppey, Ellen T. Murray, Evan R. Beiter, Andrew H. Walton, Mark W. Hall, Richard S. Hotchkiss. Comparison of monocyte human leukocyte antigen-DR expression and stimulated tumor necrosis factor alpha production as outcome predictors in severe sepsis: a prospective observational study. Critical Care. 2016;20(1):334. doi:10.1186/s13054-016-1505-0. https://link.springer.com/article/10.1186/s13054-016-1505-0 PMID:27760554; PMC5072304.

8. G. Monneret, Thomas Lafon, M. Gossez, et al.. Monocyte HLA-DR expression in septic shock patients: insights from a 20-year real-world cohort of 1023 cases. Intensive Care Medicine. 2025;51(10):1820–1832. doi:10.1007/s00134-025-08110-w. 10.1007/s00134-025-08110-w PMID:40986015; PMC12504386.

9. Martin MD, Badovinac VP, Griffith TS. CD4 T Cell Responses and the Sepsis-Induced Immunoparalysis State. Frontiers in immunology. 2020;11:1364. doi:10.3389/fimmu.2020.01364. https://pmc.ncbi.nlm.nih.gov/articles/PMC7358556/ PMID:32733454; PMC7358556.

10. de Pablo R, Monserrat J, Prieto A, Alvarez-Mon M. Role of Circulating Lymphocytes in Patients with Sepsis. BioMed research international. 2014;2014:671087. doi:10.1155/2014/671087. https://pmc.ncbi.nlm.nih.gov/articles/PMC4163419/ PMID:25302303; PMC4163419.

11. Drewry AM, Samra N, Skrupky LP, Fuller BM, Compton SM, Hotchkiss RS. Persistent lymphopenia after diagnosis of sepsis predicts mortality. Shock. 2014;42(5):383–91. doi:10.1097/SHK.0000000000000234. 10.1097/SHK.0000000000000234 PMID:25051284; PMC4362626.

12. M. Bodinier, E. Peronnet, J.-F. Llitjos, et al.. Integrated clustering of multiple immune marker trajectories reveals different immunotypes in severely injured patients. Critical Care. 2024;28(1):240. doi:10.1186/s13054-024-04990-4. 10.1186/s13054-024-04990-4 PMID:39010113; PMC11247757.

13. Dominik Schaack, Benedikt Hermann Siegler, Sandra Tamulyte, Markus Alexander Weigand, Florian Uhle. The immunosuppressive face of sepsis early on intensive care unit—A large-scale microarray meta-analysis. PLOS ONE. 2018;13(6):e0198555. doi:10.1371/journal.pone.0198555. https://journals.plos.org/plosone/article/file?id=10.1371/journal.pone.0198555&type=printable PMID:29920518; PMC6007920.

14. Andrew J. Kwok, Alice Allcock, Ricardo C. Ferreira, Eddie Cano-Gamez, Madeleine Smee, Katie L. Burnham, Yasemin-Xiomara Zurke, Stuart McKechnie, Alexander J. Mentzer, Claudia Monaco, Irina A. Udalova, Charles J. Hinds, John A. Todd, Emma E. Davenport, Julian C. Knight. Neutrophils and emergency granulopoiesis drive immune suppression and an extreme response endotype during sepsis. Nature Immunology. 2023;24(5):767–779. doi:10.1038/s41590-023-01490-5. https://www.nature.com/articles/s41590-023-01490-5 PMID:37095375.

15. J. Demaret, F. Venet, A. Friggeri, et al.. Marked alterations of neutrophil functions during sepsis-induced immunosuppression. Journal of Leukocyte Biology. 2015;98(6):1081–1090. doi:10.1189/jlb.4a0415-168rr. 10.1189/jlb.4a0415-168rr PMID:26224052.

16. Sang Ook Ha, Sang Hyuk Park, So Hee Park, Jae-Seok Park, Jin Won Huh, Chae-Man Lim, et al.. Fraction of immature granulocytes reflects severity but not mortality in sepsis. Scandinavian Journal of Clinical and Laboratory Investigation. 2015;75(1):36–43. doi:10.3109/00365513.2014.965736. 10.3109/00365513.2014.965736 PMID:25342241.

17. Ila Joshi, Walter P. Carney, Edwin P. Rock. Utility of monocyte HLA-DR and rationale for therapeutic GM-CSF in sepsis immunoparalysis. Frontiers in Immunology. 2023;14:1130214. doi:10.3389/fimmu.2023.1130214. https://www.frontiersin.org/journals/immunology/articles/10.3389/fimmu.2023.1130214/full PMID:36825018; PMC9942705.

18. Marie-Angélique Cazalis, A. Lepape, F. Venet, et al.. Early and dynamic changes in gene expression in septic shock patients: a genome-wide approach. Intensive Care Medicine Experimental. 2014;2(1):20. doi:10.1186/s40635-014-0020-3. 10.1186/s40635-014-0020-3 PMID:26215705; PMC4512996.

19. Chen D, Zhou K, Tian R, Wang R, Zhou Z. Predictive value of the dynamics of absolute lymphocyte counts for 90-day mortality in ICU sepsis patients: a retrospective big data study. BMJ Open. 2024;14(7):e084562. doi:10.1136/bmjopen-2024-084562. 10.1136/bmjopen-2024-084562 PMID:38960455; PMC11227848.

20. Fratea A, Riza AL, Dumitrescu F, Dorobantu S, Pirvu A, Dragos A, Grigorescu A, Streata I, Netea MG, Kumar V, Boahen CK. Integrated transcriptomic and proteomic profiling identifies an interferon-dependent inflammatory endotype in sepsis. Biomed Pharmacother. 2026;195:119014. doi:10.1016/j.biopha.2026.119014. 10.1016/j.biopha.2026.119014 PMID:41558266.

21. Anwen Huang, Jinxiu Wu, Jiakuan Wang, Chengwen Jiao, Yunfei Yang, Huaiwen Xiao, Li Yao. Immune gene features and prognosis in colorectal cancer: insights from ssGSEA typing. Discover Oncology. 2025;16(1):139. doi:10.1007/s12672-025-01928-2. https://link.springer.com/article/10.1007/s12672-025-01928-2 PMID:39921789; PMC11807041.

22. Yukari Kobayashi, Yoshihiro Kushihara, Noriyuki Saito, et al.. A novel scoring method based on RNA-Seq immunograms describing individual cancer-immunity interactions. Cancer Science. 2020;111(11):4031–4040. doi:10.1111/cas.14621. 10.1111/cas.14621 PMID:32810311; PMC7648030.

23. Barbie DA, Tamayo P, Boehm JS, Kim SY, Moody SE, Dunn IF, Schinzel AC, Sandy P, Meylan E, Scholl C, Fröhling S, Chan EM, Sos ML, Michel K, Mermel C, Silver SJ, Weir BA, Reiling JH, Sheng Q, Gupta PB, Wadlow RC, Le H, Hoersch S, Wittner BS, Ramaswamy S, Livingston DM, Sabatini DM, Meyerson M, Thomas RK, Lander ES, Mesirov JP, Root DE, Gilliland DG, Jacks T, Hahn WC. Systematic RNA interference reveals that oncogenic KRAS-driven cancers require TBK1. Nature. 2009;462(7269):108–12. doi:10.1038/nature08460. 10.1038/nature08460 PMID:19847166; PMC2783335.

24. Hänzelmann S, Castelo R, Guinney J. GSVA: gene set variation analysis for microarray and RNA-seq data. BMC Bioinformatics. 2013;14:7. doi:10.1186/1471-2105-14-7. 10.1186/1471-2105-14-7 PMID:23323831; PMC3618321.

25. Alexander T. Wenzel, John Jun, Pablo Tamayo, Jill P. Mesirov. Profiling ranked list enrichment scoring in sparse data elucidates algorithmic tradeoffs. bioRxiv (preprint; not peer-reviewed). 2024. Preprint — no volume/issue/pages. doi:10.1101/2024.06.03.597180. 10.1101/2024.06.03.597180

26. Courtney Bull, Ryan M Byrne, Natalie C Fisher, Shania M Corry, Raheleh Amirkhah, Jessica Edwards, Lily Hillson, Mark Lawler, Aideen Ryan, Felicity Lamrock, Philip D Dunne, Sudhir B Malla. Evaluation of Gene Set Enrichment Analysis (GSEA) tools highlights the value of single sample approaches over pairwise for robust biological discovery. bioRxiv (preprint; not peer-reviewed). 2024. Preprint — no volume/issue/pages. doi:10.1101/2024.03.15.585228. https://www.biorxiv.org/content/10.1101/2024.03.15.585228v1

27. Mohammad Alì Mansournia, Angelika Geroldinger, Sander Greenland, Georg Heinze. Separation in Logistic Regression: Causes, Consequences, and Control. American Journal of Epidemiology. 2018;187(4):864–870. doi:10.1093/aje/kwx299. 10.1093/aje/kwx299 PMID:29020135.

28. Heinze G, Schemper M. A solution to the problem of separation in logistic regression. Statistics in Medicine. 2002;21(16):2409–2419. doi:10.1002/sim.1047. https://onlinelibrary.wiley.com/doi/10.1002/sim.1047 PMID:12210625.

29. R. Puhr, G. Heinze, Mariana Nold, et al.. Firth’s logistic regression with rare events: accurate effect estimates and predictions?. Statistics in Medicine. 2017;36(14):2302–2317. doi:10.1002/sim.7273. 10.1002/sim.7273 PMID:28295456.

30. Georg Heinze. A comparative investigation of methods for logistic regression with separated or nearly separated data. Statistics in Medicine. 2006;25(24):4216–4226. doi:10.1002/sim.2687. 10.1002/sim.2687 PMID:16955543.

31. Jamie O. Yang, M. Zinter, M. Pellegrini, et al.. Whole blood transcriptomics identifies subclasses of pediatric septic shock. Critical Care. 2023;27(1):486. doi:10.1186/s13054-023-04689-y. 10.1186/s13054-023-04689-y PMID:38066613; PMC10709863.

32. Francois B, Jeannet R, Daix T, Walton AH, Shotwell MS, Unsinger J, Monneret G, Rimmelé T, Blood T, Morre M, Gregoire A, Mayo GA, Blood J, Durum SK, Sherwood ER, Hotchkiss RS. Interleukin-7 restores lymphocytes in septic shock: the IRIS-7 randomized clinical trial. JCI Insight. 2018;3(5). doi:10.1172/jci.insight.98960. 10.1172/jci.insight.98960 PMID:29515037; PMC5922293.

33. Li D, Zhang J, Cheng W, Zhao G, Lei X, Xie Y, Cui N, Wang H. Dynamic changes in peripheral blood lymphocyte trajectory predict the clinical outcomes of sepsis. Frontiers in Immunology. 2025;16:1431066. doi:10.3389/fimmu.2025.1431066. 10.3389/fimmu.2025.1431066 PMID:39967662; *PMC11832464*.

